# Optimal Donor Selection Across Multiple Outcomes For Hematopoietic Stem Cell Transplantation By Bayesian Nonparametric Machine Learning

**DOI:** 10.1101/2024.05.09.24307134

**Authors:** Rodney A Sparapani, Martin Maiers, Stephen R. Spellman, Bronwen E Shaw, Purushottam W Laud, Steven M. Devine, Brent R Logan

## Abstract

Allogeneic hematopoietic cell transplantation (HCT) is one of the only curative treatment options for patients suffering from life-threatening hematologic malignancies; yet, the possible adverse complications can be serious even fatal. Matching between donor and recipient for 4 of the HLA genes is widely accepted and supported by the literature. However, among 8/8 allele matched unrelated donors, there is less agreement among centers and transplant physicians about how to prioritize donor characteristics like additional HLA loci (DPB1 and DQB1), donor sex/parity, CMV status, and age to optimize transplant outcomes. This leads to varying donor selection practice from patient to patient or via center protocols. Furthermore, different donor characteristics may impact different post transplant outcomes beyond mortality, including disease relapse, graft failure/rejection, and chronic graft-versus-host disease (components of event-free survival, EFS). We develop a general methodology to identify optimal treatment decisions by considering the trade-offs on multiple outcomes modeled using Bayesian nonparametric machine learning. We apply the proposed approach to the problem of donor selection to optimize overall survival and event-free survival, using a large outcomes registry of HCT recipients and their actual and potential donors from the Center for International Blood and Marrow Transplant Research (CIBMTR). Our approach leads to a donor selection strategy that favors the youngest male donor, except when there is a female donor that is substantially younger.

## 1 Introduction

The discovery of stem cells in 1963 [McCulloch and Till, 2005] has lead to new avenues of treatment for a variety of illnesses including blood-borne cancers, auto-immune diseases and inherited newborn conditions. Allogeneic hematopoietic cell transplantation (HCT) is the only curative treatment option for patients suffering from life-threatening hematologic malignancies; yet, the possible adverse complications can be serious even fatal. Stem cells are harvested from a donor that is HLA matched to the transplant recipient. In 2021 for the United States (US) alone, 9,349 allogeneic HSCT treatments were performed [Center for International Blood and Marrow Transplant Research, 2024] and unrelated volunteers donated their stem cells for 5,073 (54%) of those (the remainder of donors were kin). The National Marrow Donor Program (NMDP) in the US maintains the world’s largest stem cell registry of more than 9 million donors and potential donors, and facilitates access to more then 42 million donors world-wide through the World Marrow Donor Association. For patients without an HLA-matched family member, a donor search is conducted by the NMDP registry. HLA matching is critical to preventing/mitigating graft vs. host disease (GVHD) by ensuring antigen cross-compatibility with the transplanted immune system. However, all matched unrelated donors (MUD) do not necessarily have the same prognostic risks/benefits. Besides HLA loci, other donor characteristics are considered including age, sex/child-bearing parity, cytomegalovirus (CMV) serostatus. Previous research has consistently found survival benefits associated with choosing a younger MUD [Kollman et al., 2001, 2016, Shaw et al., 2018, Pidala et al., 2019, Logan et al., 2021]. But, the optimization potential for selection of other donor factors has not been consistently shown [Shaw et al., 2007, Fleischhauer et al., 2012, Pidala et al., 2014, Fleischhauer et al., 2017, Shaw et al., 2017]. Therefore, donor selection practices vary from patient to patient (or via center to center protocols), providing us with the opportunity to utilize modern machine learning techniques to determine which factors will yield the best possible outcomes.

Several complications arise in optimizing donor selection. First, donor selection requires consideration of multiple donor factors from a finite but sometimes large list of potential MUDs specific to a given recipient. Therefore, practical implementation of optimal donor selection requires a good understanding of what donor characteristics are necessary to be considered in an optimization algorithm. Second, donor selection should be individualized, since the impact of donor factors may be dependent on patient or disease characteristics; this necessitates that a prediction model for patient outcomes has sufficient flexibility to capture complex interactions. Third, selecting an optimal donor depends on what outcome is being optimized. Overall survival is of greatest importance, but there are other post-transplant complications such as clinical relapse, graft failure and/or GVHD causing severe morbidity that could be impacted by donor characteristics.

We develop optimal donor selection methodology as an individualized decision for a potential recipient while considering multiple outcomes along with the likely trade-offs amongst them. Prior to optimization, we examine the impact of various donor characteristics on relevant outcomes: narrowing to a relatively parsimonious subset for the implementation of the donor search. Next, we develop an optimal donor selection method with multiple outcomes. Qian and Murphy [2011] show that the optimal individualized treatment rule assigns each patient to the treatment with the optimal conditional expectation given their patient characteristics. Following this, we propose to optimize donor selection by directly selecting the donor whose characteristics optimize the expected outcome for a given patient based on a prediction model. Furthermore, accurate predictive modeling is sufficient to identify an optimal treatment rule [Qian and Murphy, 2011]. From training data, we construct prediction models for overall survival (OS) and event-free survival (EFS is a composite of death, clinical relapse, graft failure/rejection or moderate/severe chronic GVHD) with a non-parametric machine learning framework based on Bayesian Additive Regression Trees (BART) Chipman et al. [2010]. BART models have three key features which make them useful in this setting: 1) excellent predictive performance; 2) automatically incorporate complex interactions; and 3) avoiding precarious restrictive assumptions like linearity. On a solid foundation of Bayesian inference, BART inherently provides uncertainty quantification of any model predictions as well as any function of them. The development of BART models for survival outcomes has demonstrated increasing flexibility [Bonato et al., 2011, Sparapani et al., 2016, Henderson et al., 2020, Linero et al., 2022, Sparapani et al., 2023]. We will employ Nonparametric Failure Time BART (NFT BART) [Sparapani et al., 2023] for predictive modeling that avoids restrictive assumptions (such as proportionality, homoskedasticity and normality) while providing computational scalability to large data sets like we have here. After predictive modeling is performed for each outcome, we create a weighted utility from the OS and EFS expectations from each potential donor to a given recipient. Now, we select the donor which optimizes the weighted utility function. Weights represent the desired relative importance of the outcomes. We demonstrate several optimal donor selection policies via different weights.

## 2 Methods

### 2.1 Data Sources

Clinical outcome data was obtained from the Center for International Blood and Marrow Transplant Research (CIBMTR) research database. CIBMTR is a research collaboration between the NMDP and the Medical College of Wisconsin which collects outcome data for all allogeneic HSCT recipients in the US as the custodian of the Stem Cell Therapeutic Outcomes Database under the C.W. Bill Young Transplantation Program of the Health Resources and Services Administration [Stem Cell Therapeutic and Research Act Reauthorization, 2021]. Our cohort consisted of all HCT recipients in the US from 2016 to 2019 with 8/8 high-resolution matching at HLA-A, B, C, and DRB1 to their unrelated donor. All patients provided informed consent for participation in the CIBMTR Research Database and the study was approved by the NMDP Institutional Review Board. We randomly divided our cohort into a subset of 10,016 patients (85%) for training prediction models and 1,802 (15%) for validation of the prediction models. Within the validation subset, 699 (39%) had their search archive records available. The NMDP search archive database is a snapshot of each patient’s donor search prior to transplant that includes all of the potentially matched unrelated donors on the registry at the time of the search. This allows us to re-conduct the donor search using our proposed donor selection algorithms and assess how the proposed strategies will perform in practice, compared to the real-world donor selection practice based on the actual donor selected. Since we typically do not know the high-resolution HLA typing and match status of all potential donors at the time of the search, we restrict the donor list for each patient in the search archive subset to likely 8/8 matches (for volunteers whose HLA typing is ambiguous [Paunić et al., 2016], we only consider those matches with a probability ≥ 0.9 based on HapLogic predictions [Dehn et al., 2016]). The additional donor characteristics beyond the requisite 8/8 HLA matching to be considered for optimal selection are age, sex/child-bearing parity, CMV status, HLA -DPB1 and/or -DQB1.

### 2.2 Study Endpoints

We focus on optimizing both overall survival/mortality (OS) and event-free survival (EFS) that is defined as a composite of death, clinical relapse, graft failure/rejection, moderate/severe chronic GVHD: whichever comes first. Since transplant is a curative therapy with OS/EFS curves flattening substantially by 3 years, we focus on this time horizon by optimizing OS/EFS by either the point-wise 3 year probabilities, or the restricted mean survival time (RMST) up to 3 years. RMST is an alternative framing of survival outcomes that may have advantages over point-wise analysis or proportional hazards modeling, particularly in interpretation [Royston and Parmar, 2013, Pak et al., 2017, Kloecker et al., 2020].

### 2.3 Statistical Analysis

We fit NFT BART prediction models to the OS and EFS data using the **nft-bart** R package [Sparapani et al., 2023]. Posterior samples of survival predictions are generated for a patient *p* with characteristics *x*_*p*_ who is a recipient of transplantation from donor *d* with characteristics *z*_*d*_, given by *S*_*m*_(*t*|*x*_*p*_, *z*_*d*_, 𝒟), for draws *m* = 1, …, *M*. Here we use 𝒟 in this Bayesian model to represent that inference in the model is conditional on the observed data, including the event/censoring times, event indicators, and measured covariates. Similarly, we generate posterior samples of predictions for RSMT up to time *t*, defined by RMST 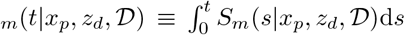 (see Section 6 of the Supplement for more details on these calculations). We use the posterior mean of these samples to summarize predictions and to perform optimizations; in particular, *E*[*S*(*t*|*x*_*p*_, *z*_*d*_, 𝒟)] ≈ *M* ^−1^ ∑_*m*_*S*_*m*_(*t*|*x*_*p*_, *z*_*d*_, 𝒟) and *E*[RMST(*t*|*x*_*p*_, *z*_*d*_, 𝒟)] ≈*M* ^−1^ ∑_*m*_RMST_*m*_(*t*|*x*_*p*_,*z*_*d*_, 𝒟). Furthermore, the posterior samples can be used to quantify uncertainty, e.g., the (1 − *q*) × 100% credible interval for survival is *S*_*m*_(*t*|*x*_*p*_, *z*_*d*_, 𝒟) : *S*_*m*_*t* (*t*|*x*_*p*_, *z*_*d*_, 𝒟) where *m* (*m*^*′*^) is the *q/*2 × 100% (1 − *q/*2 × 100%) posterior quantile. Waterfall plots [Gillespie, 2012] were generated to describe changes in predicted outcomes for each patient as an individual donor characteristic was varied one at a time while holding the others fixed. For example, the waterfall plot for donor sex in the bottom row of Figure 1 shows the predictions (posterior means) for each transplant if the donor had been male vs. female. Waterfall plots showing little to no impact in survival for all, or nearly all, patients attributable to a donor characteristic change indicate a negligible impact that can be simply ignored. We define a negligible difference of *<* 1 % in predicted survival at 3 years or *<* 10 days in RMST as an indifference zone [Soeteman et al., 2020]. After this donor characteristic selection was completed the NFT BART model was refitted with the relevant donor characteristics for further evaluation on donor optimization. Note that prediction models are built for both OS and EFS. Notationally, we refer to the posterior mean OS or EFS as *E*[OS(*t*|*x*_*p*_, *z*_*d*_, 𝒟] and *E*[EFS(*t*|*x*_*p*_, *z*_*d*_, 𝒟] respectively; and to posterior mean RMST OS or EFS as *E*[RMOS(*t*|*x*_*p*_, *z*_*d*_, 𝒟)] and *E*[RMEFS(*t*|*x*_*p*_, *z*_*d*_, 𝒟)] respectively.

**Figure 1:**
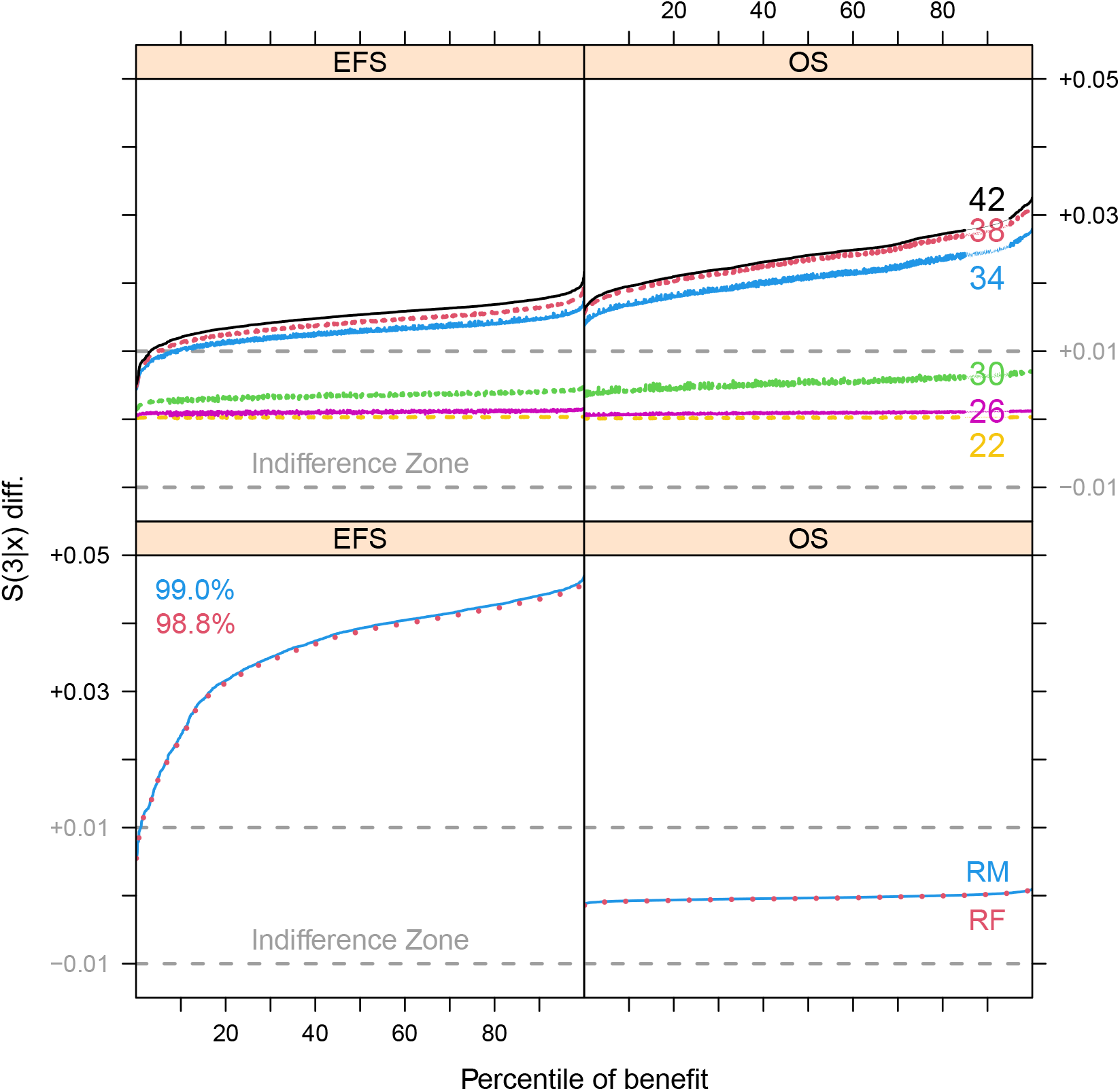
Waterfall plots of EFS and OS differentials based on predictions for the validation set. In the first (second) column, we have EFS (OS) differentials on the vertical axis, with an Indifference Zone in grey, and percentile of benefit on the horizontal axis. In the top row, differentials for an older donor vs. an 18 year-old; donor ages (lines) in ascending sequence: 22 (dashed yellow), 26 (solid magenta), 30 (dashed green), 34 (solid blue), 38 (dashed red) and 42 (solid black). In the bottom row, differentials for a male donor vs. a female; recipient male (female) with a solid blue line (dotted red line): in the left panel, the percentage of those that benefit from a male vs. female donor for recipient males (females) in blue (red).

To optimize donor selection, we follow the approach of Qian and Murphy [2011] who show that an optimal individualized treatment rule assigns each patient to the treatment which has the best conditional expectation given their patient characteristics. Here we define an optimal donor selection rule by selecting the donor which has the best expected outcome from our NFT BART prediction model according to the selected donor features. To address multiple outcomes of OS and EFS, we optimize a utility function which is a weighted average of the OS and EFS outcomes. The weight parameter represents the relative importance of the corresponding outcome in terms of the donor selection rule. The optimal donor for patient *p* among their set *D*_*p*_ of potential donors is defined as

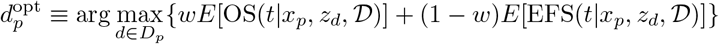

for the pointwise survival probability outcome (and similarly defined for RMST). Note that a weight of *w* = 1 represents donor optimization based solely on OS as opposed to *w* = 0 for EFS only. Weights between 0.5 *< w <* 1 tend to have greater emphasis on optimizing OS, yet still improving EFS particularly in situations where OS differences are small. The posterior mean OS and EFS probabilities for this optimal donor are 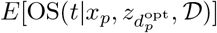 and 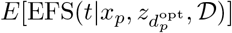 respectively.

We benchmark the optimal donor strategy performance against the actual donor 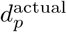 used for each transplant in the search archive by taking the difference in posterior mean OS or EFS, according to the following.

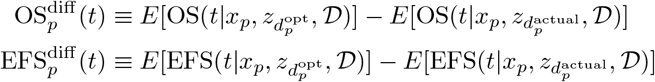

Plots of 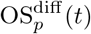 vs. 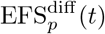 by patient for different values of *w* are constructed to show how *w* impacts the tradeoffs between the two outcomes.

The population level outcome for an optimal donor strategy is obtained by averaging the optimal outcomes across a sample of patients as follows.

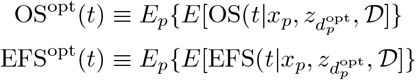

Population level outcomes for the actual donor strategy can be similarly defined, as well as differences in population level outcomes between the optimal and actual donor strategy. Note also that posterior samples of the population level outcomes can be obtained by applying the optimization on a posterior sample basis. Finally, we have shown an optimal donor selection rule based on the weighted average of the OS and EFS probabilities at time *t*. An optimal donor selection rule based on a weighted average of the RMST for OS and EFS up to time *t* could be similarly derived.

## 3 Results

Among the recipients within the training (validation) set, 37.3% (38.4%) died whereas among the censored survivors the median days of follow-up was 749 (745) with a first:third quartile of 391:1117 (390:1110). Similarly, for EFS, 61.3% (61.6%) suffered the event with median survivor days of follow-up of 741 (737) and first:third quartile 383:1107 (382:1100) for training (validation) respectively. We summarize the collected data in a series of tables for recipients, donors and disease characteristics: the **bold** variables are included in the model (additional model variables that are not shown here appear in the Supplement Tables 2 through 6). The cohort of recipients consists mainly of those 40 or older: about three-quarters (74.1%); see Table 1. The donors are comparatively younger: 88.5% are below 40; see Table 2. Almost three-quarters of the patients, 73%, were stricken by the three most common hematologic cancers: acute lymphoblastic leukemia (ALL), 13%; acute myeloid leukemia (AML), 40%; and myelodysplastic syndrome (MDS), 20%; see Table 3.

**Table 1:**
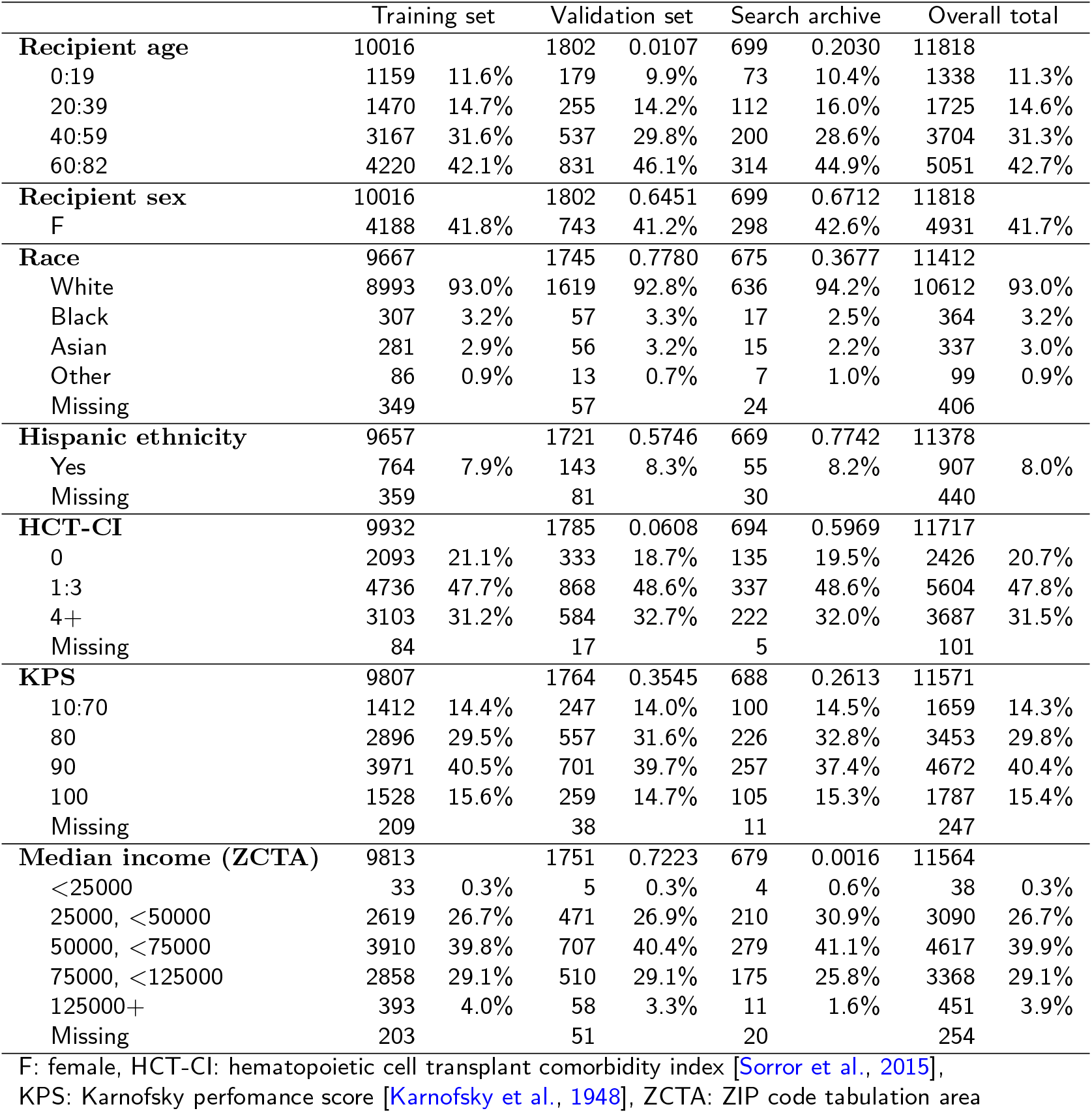
Recipient demographic characteristics. Training and validation sets are mutually exclusive while search archive is a subset of validation. Within the total rows, we present Pearson’s Chi-squared test p-values for the comparisons between training with validation and training with search archive: missing values excluded. **Bold** variables are included in the model.

**Table 2:** Donor matching characteristics. Training and validation sets are mutually exclusive while search archive is a subset of validation. Within the total rows, we present Pearson’s Chi-squared test p-values for the comparisons between training with validation and training with search archive: missing values excluded. **Bold** variables are included in the model.

|  | Training set |  | Validation set |  | Search archive |  | Overall total |  |
| --- | --- | --- | --- | --- | --- | --- | --- | --- |
| **Donor age** | 9941 |  | 1791 | 0.9524 | 698 | 0.3400 | 11732 |  |
| 17:29 | 6499 | 65.4% | 1176 | 65.7% | 479 | 68.6% | 7675 | 65.4% |
| 30:39 | 2291 | 23.0% | 411 | 22.9% | 146 | 20.9% | 2702 | 23.0% |
| 40:49 | 855 | 8.6% | 148 | 8.3% | 52 | 7.4% | 1003 | 8.5% |
| 50:60 | 296 | 3.0% | 56 | 3.1% | 21 | 3.0% | 352 | 3.0% |
| Missing | 75 |  | 11 |  | 1 |  | 86 |  |
| **Donor sex** | 9957 |  | 1794 | 0.5212 | 698 | 0.0059 | 11751 |  |
| M | 6985 | 70.2% | 1245 | 69.4% | 524 | 75.1% | 8230 | 70.0% |
| Missing | 59 |  | 8 |  | 1 |  | 67 |  |
| Donor parity | 9957 |  | 1794 | 0.1666 | 698 | <0.0001 | 11751 |  |
| M | 6985 | 70.2% | 1245 | 69.4% | 524 | 75.1% | 8230 | 70.0% |
| F Nonparous | 1886 | 18.9% | 370 | 20.6% | 137 | 19.6% | 2256 | 19.2% |
| F Parous | 1086 | 10.9% | 179 | 10.0% | 37 | 5.3% | 1265 | 10.8% |
| Missing | 59 |  | 8 |  | 1 |  | 67 |  |
| Sex match | 9957 |  | 1794 | 0.8539 | 698 | 0.7195 | 11751 |  |
| Yes | 5751 | 57.8% | 1032 | 57.5% | 408 | 58.5% | 6783 | 57.7% |
| Missing | 59 |  | 8 |  | 1 |  | 67 |  |
| CMV match | 9962 |  | 1786 | 0.7653 | 691 | 0.4890 | 11748 |  |
| Yes | 5560 | 55.8% | 990 | 55.4% | 395 | 57.2% | 6550 | 55.8% |
| Missing | 54 |  | 16 |  | 8 |  | 70 |  |
| DPB1 match | 8526 |  | 1535 | 0.4652 | 603 | 0.3261 | 10061 |  |
| M/P | 6430 | 75.4% | 1171 | 76.3% | 444 | 73.6% | 7601 | 75.5% |
| Missing | 1490 |  | 267 |  | 96 |  | 1757 |  |
| DQB1 match | 9810 |  | 1773 | 0.6492 | 689 | 0.1282 | 11583 |  |
| Yes | 9377 | 95.6% | 1699 | 95.8% | 667 | 96.8% | 11076 | 95.6% |
| Missing | 206 |  | 29 |  | 10 |  | 235 |  |

CMV: cytomegalovirus, F: female, M: male,
M/P: either a match or a permissive mismatch [Zino et al., 2004]

**Table 3:** Disease and transplant characteristics. Training and validation sets are mutually exclusive while search archive is a subset of validation. Within the total rows, we present Pearson’s Chi-squared test p-values for the comparisons between training with validation and training with search archive: missing values excluded. **Bold** variables are included in the model.

|  | Training set |  | Validation set |  | Search archive |  | Overall total |  |
| --- | --- | --- | --- | --- | --- | --- | --- | --- |
| **Disease** | 10016 |  | 1802 | 0.0380 | 699 | 0.1416 | 11818 |  |
| ALL | 1290 | 12.9% | 251 | 13.9% | 102 | 14.6% | 1541 | 13.0% |
| AML | 3956 | 39.5% | 746 | 41.4% | 290 | 41.5% | 4702 | 39.8% |
| MDS | 1988 | 19.8% | 362 | 20.1% | 139 | 19.9% | 2350 | 19.9% |
| Other | 2782 | 27.8% | 443 | 24.6% | 168 | 24.0% | 3225 | 27.3% |
| **ALL/AML/MDS status** | 6947 |  | 1306 | 0.9271 | 505 | 0.0398 | 8253 |  |
| Early | 4159 | 59.9% | 775 | 59.3% | 279 | 55.2% | 4934 | 59.8% |
| Intermediate | 1360 | 19.6% | 261 | 20.0% | 99 | 19.6% | 1621 | 19.6% |
| Advanced | 1428 | 20.6% | 270 | 20.7% | 127 | 25.1% | 1698 | 20.6% |
| Missing | 287 |  | 53 |  | 26 |  | 340 |  |
| **Graft type** | 10016 |  | 1802 | 0.3259 | 699 | 0.8784 | 11818 |  |
| Bone marrow | 2239 | 22.4% | 384 | 21.3% | 158 | 22.6% | 2623 | 22.2% |
| Peripheral blood | 7777 | 77.6% | 1418 | 78.7% | 541 | 77.4% | 9195 | 77.8% |
| **Total body irradiation** | 9925 |  | 1781 | 0.9647 | 695 | 0.3402 | 11706 |  |
| Yes | 2397 | 24.2% | 431 | 24.2% | 179 | 25.8% | 2828 | 24.2% |
| Missing | 91 |  | 21 |  | 4 |  | 112 |  |
| **Conditioning regimen** | 10016 |  | 1802 | 0.7367 | 699 | 0.4458 | 11818 |  |
| Myeloablative | 4974 | 49.7% | 885 | 49.1% | 364 | 52.1% | 5859 | 49.6% |
| Non-myeloablative | 1122 | 11.2% | 213 | 11.8% | 77 | 11.0% | 1335 | 11.3% |
| Reduced intensity | 3920 | 39.1% | 704 | 39.1% | 258 | 36.9% | 4624 | 39.1% |
| **GVHD prophylaxis** | 10016 |  | 1802 | 0.0676 | 699 | 0.0026 | 11818 |  |
| CSA+MMF | 430 | 4.3% | 84 | 4.7% | 40 | 5.7% | 514 | 4.3% |
| CSA+MTX | 356 | 3.6% | 61 | 3.4% | 30 | 4.3% | 417 | 3.5% |
| CSA only | 65 | 0.6% | 14 | 0.8% | 7 | 1.0% | 79 | 0.7% |
| Cyclophosphamide | 1130 | 11.3% | 216 | 12.0% | 49 | 7.0% | 1346 | 11.4% |
| FK506+MMF | 1097 | 11.0% | 209 | 11.6% | 89 | 12.7% | 1306 | 11.1% |
| FK506+MTX | 5425 | 54.2% | 965 | 53.6% | 377 | 53.9% | 6390 | 54.1% |
| FK506 only | 1020 | 10.2% | 196 | 10.9% | 82 | 11.7% | 1216 | 10.3% |
| Other | 493 | 4.9% | 57 | 3.2% | 25 | 3.6% | 550 | 4.7% |
| **Year of transplant** | 10016 |  | 1802 | 0.3262 | 699 | <0.0001 | 11818 |  |
| 2016 | 2347 | 23.4% | 419 | 23.3% | 336 | 48.1% | 2766 | 23.4% |
| 2017 | 2527 | 25.2% | 438 | 24.3% | 324 | 46.4% | 2965 | 25.1% |
| 2018 | 2593 | 25.9% | 450 | 25.0% | 33 | 4.7% | 3043 | 25.7% |
| 2019 | 2549 | 25.4% | 495 | 27.5% | 6 | 0.9% | 3044 | 25.8% |

ALL: acute lymphoblastic leukemia, AML: acute myelogenous leukemia, CSA: Cyclosporin A, FK506: tacrolimus, GVHD: graft vs. host disease, MDS: myelodysplastic syndrome, MMF: mycophenolate mofetil, MTX: methotrexate

Now we turn to the decision-making process for which donor characteristics will optimize recipient outcomes. In Supplement Table 7, you will find a list of waterfall plots representing donor choices with respect to age, sex/parity, CMV, DPB1 and DQB1. First, we eliminate those factors that are not promising. Matching based on the criteria of CMV and DPB1 was not productive for either OS or EFS: in all cases, the expected benefit is within the indifference zone; see Supplement Figures 12, 14, 16 and 18. With respect to DQB1, we can see that matching is already being undertaken at a high rate (Table 2): only 4.4% are mismatched when available. Therefore, it is possible that we did not have sufficient data to investigate matching of DQB1 due to the relatively few mismatches; waterfall plots are provided by Supplement Figures 20 and 22. Waterfall plots for parity (parous vs. non-parous females) show that the difference between these donors is negligible; see Supplement Figures 7 to 10.

This leaves us with just age and sex. So, we refit the models narrowing the donor characteristics to age and sex for optimization predicting OS and EFS outcomes for each patient in the validation set. The plot for age shows that choosing a younger donor is beneficial for OS and to a lesser extent EFS: generally, a donor aged 30 or less is preferable with marginal gains achieved going further than that; see the upper half of Figure 1. For OS, the choice of sex (male vs. female) is indifferent as seen in the bottom right of Figure 1. But, on the contrary for EFS, the choice of sex is quite important: males are generally preferable to females, with the effect of sex being larger than the donor age effect for EFS; see the bottom left of Figure 1. Putting this together for both age and sex across both EFS and OS endpoints, we see that the youngest male is generally preferable due to EFS benefit except when a much younger female is available. In the latter case the OS benefit from younger age may be more clinically important than an EFS benefit from the male donor. As we might expect, this story is largely the same if we are considering differences in RMST rather than survival probabilities; see Figure 2. In Figure 3, we see scatter-plots for OS(3) vs. EFS(3) differentials with three donor choice strategies. Also shown in the figure is the number of female and male donors selected outside and inside the indifference zone. Optimizing OS(3) favors the youngest female (355 females and 103 males outside the indifference zone). Optimizing EFS(3) favors the youngest male (2 females and 280 males outside the indifference zone). Finally, optimizing 2OS(3):1EFS(3) generally favors the youngest male except when there is a considerably younger female available (35 females and 253 males outside the indifference zone). In Figure 4, we plot the population-level value function with respect to these donor choice strategies; optimizing 2OS(3):1EFS(3) provides near-optimal performance for OS(3) and EFS(3) differentials respectively.

**Figure 2:**
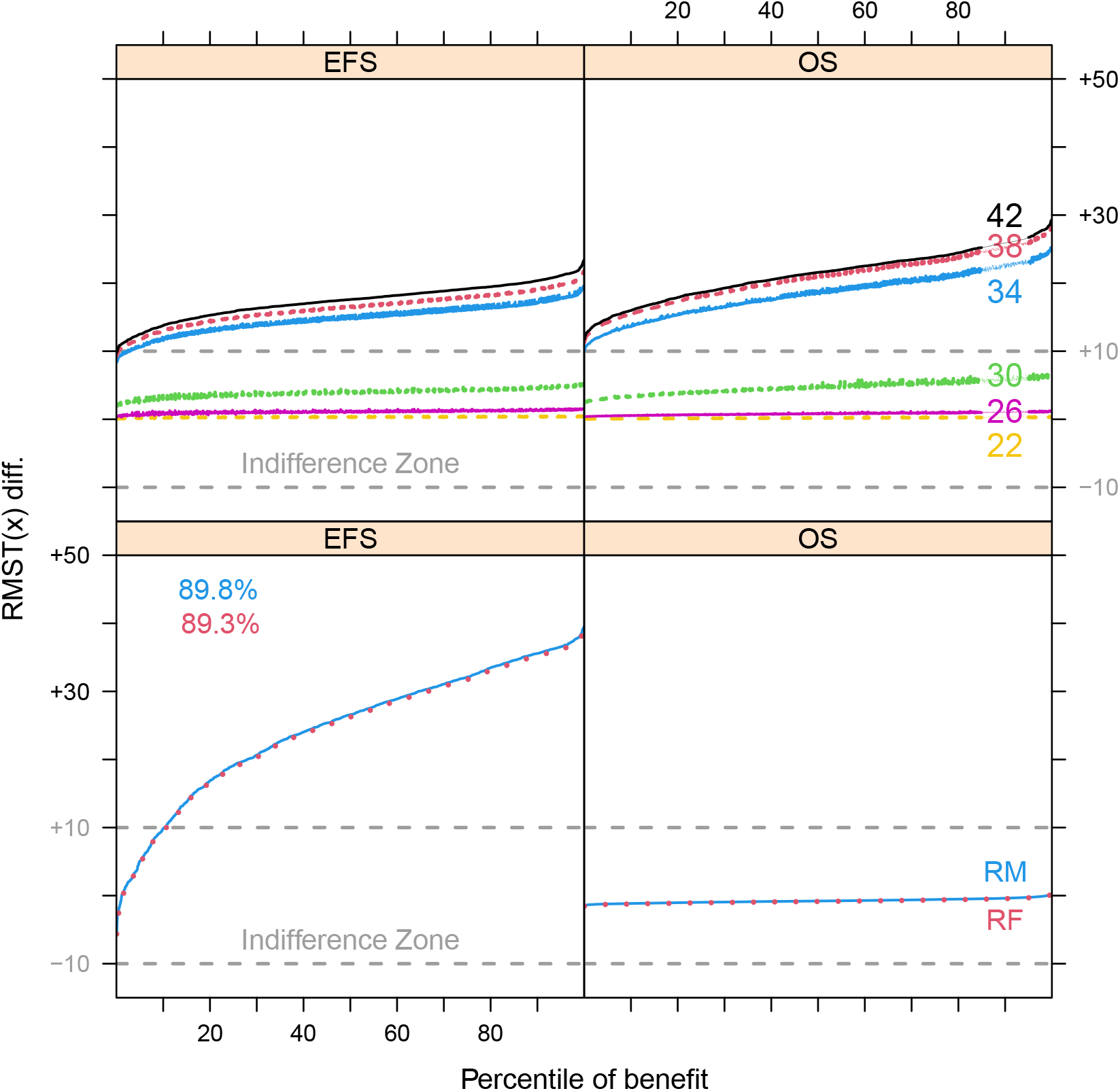
Waterfall plots of EFS and OS differentials based on RMST predictions in days for the validation set. In the first (second) column, we have EFS (OS) RMST differentials on the vertical axis, with an Indifference Zone in grey, and percentile of benefit on the horizontal axis. In the top row, differentials for an older donor vs. an 18 year-old; donor ages (lines) in ascending sequence: 22 (dashed yellow), 26 (solid magenta), 30 (dashed green), 34 (solid blue), 38 (dashed red) and 42 (solid black). In the bottom row, differentials for a male donor vs. a female; recipient male (female) with a solid blue line (dotted red line): in the left panel, the percentage of those that benefit from a male vs. female donor for recipient males (females) in blue (red).

**Figure 3:**
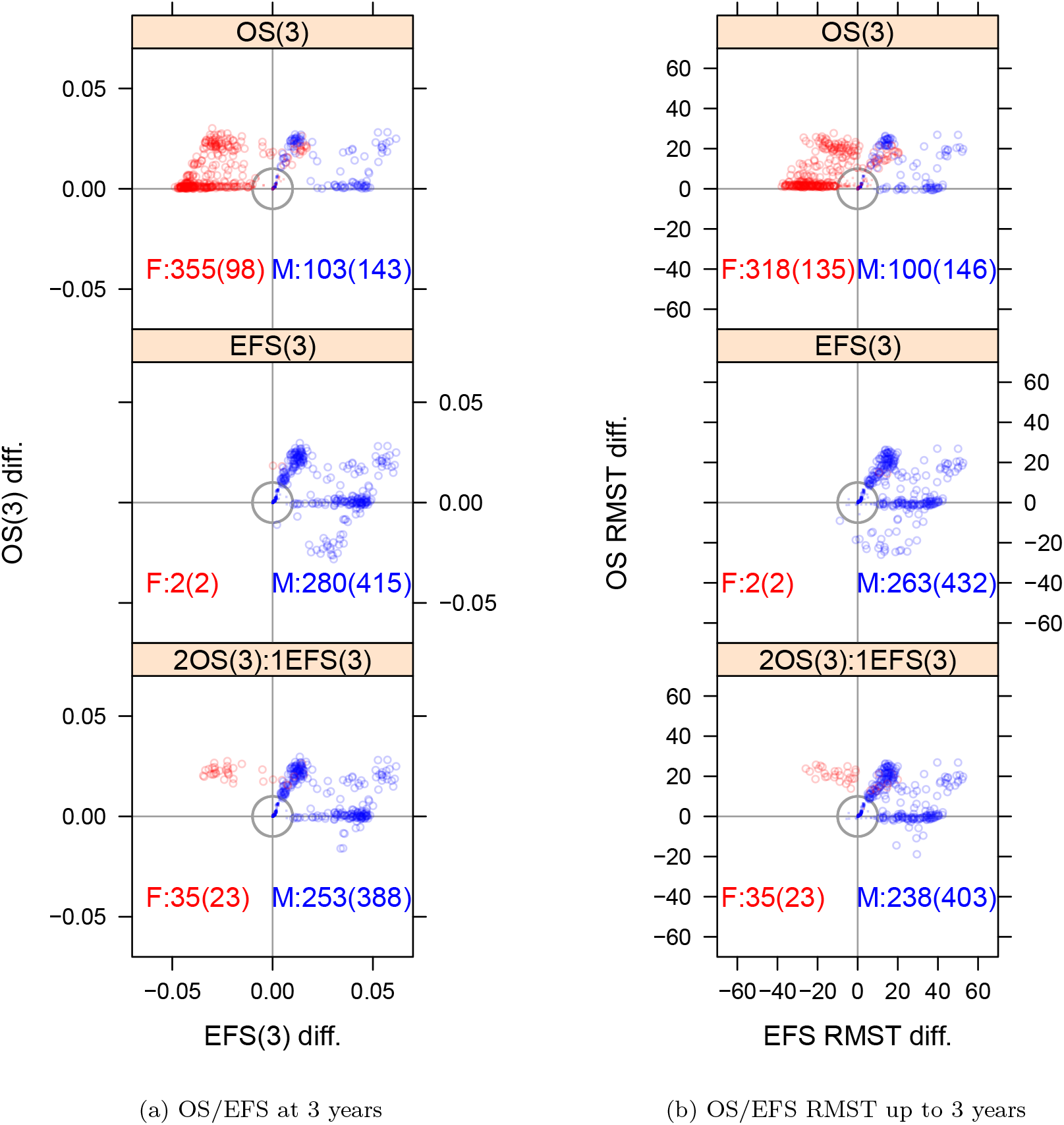
Optimal donor selection for OS and EFS among search archive recipients. In each row of the figures, optimal is determined by OS (top), EFS (middle) and weighted 2OS:1EFS (bottom) respectively for survival probability differentials at 3 years. On the *y*-axis (*x*-axis), we have OS (EFS) differentials of the optimal matching donor vs. the actual donor. At the bottom of each plot, we have a summary of the youngest female (F) donor vs. youngest male (M) chosen: beyond (within) the Indifference Zone is the first summary (second summary in parentheses). Red (blue) circles represent females (males) beyond the Indifference Zone circle boundary (grey line); red (blue) dots are females (males) within the Indifference Zone.

**Figure 4:**
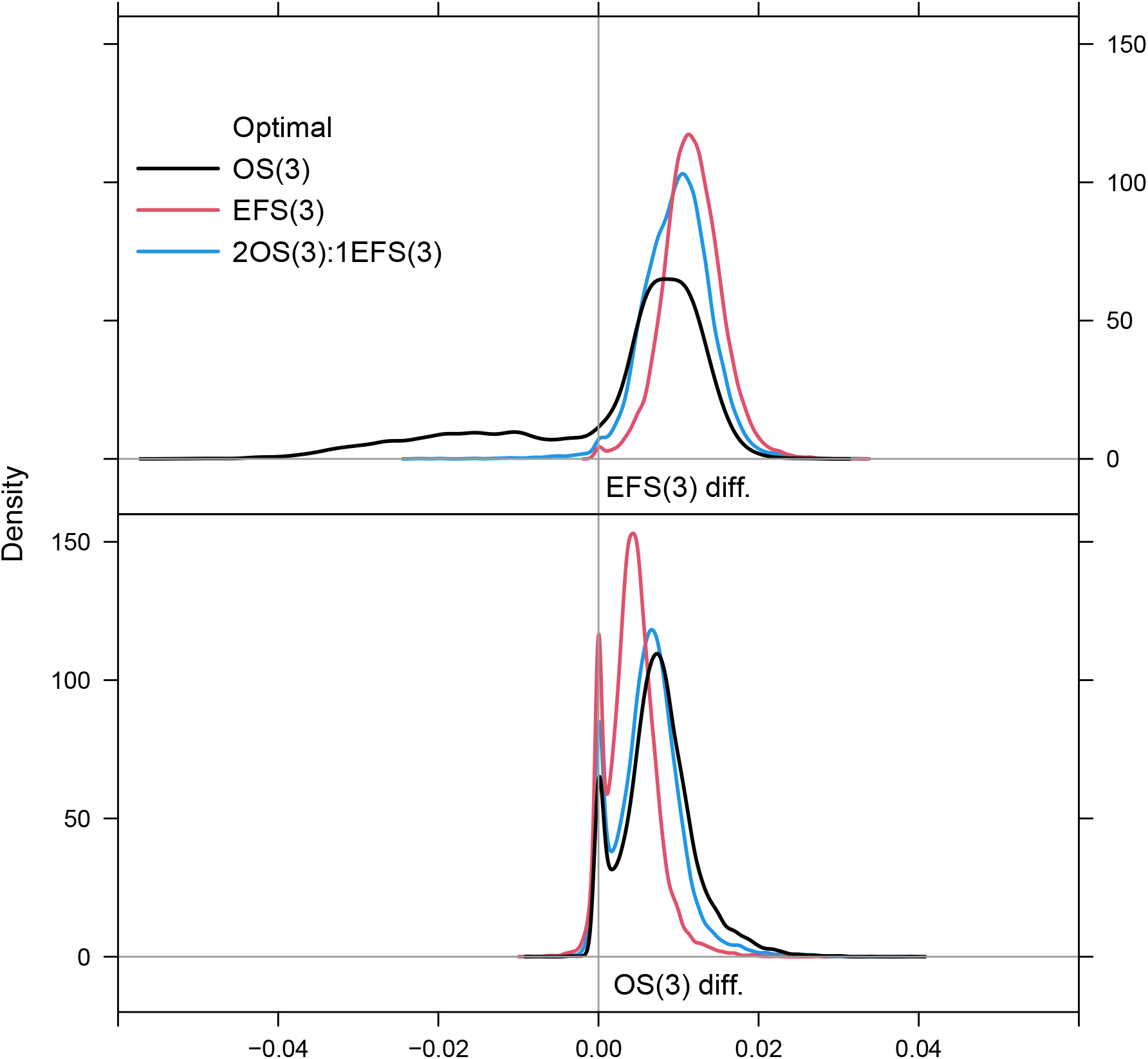
Population-level value function for optimal donor selection survival probability differentials among search archive recipients. The top (bottom) row is for EFS (OS). Three optimal strategy lines are presented for year 3 survival probability differentials: OS (black), EFS (red) and 2OS:1EFS (blue).

## 4 Conclusions

In this article, we proposed a novel approach to optimize treatment across multiple outcome variables, using a weighted utility function, to prioritize treatments having the best results for the most important clinical outcomes. We used a flexible machine learning approach for survival data called NFT BART to build prediction models. This model makes minimal assumptions, while automatically handling complex relationships with non-linearity and interactions, allowing for patient specific predictions of survival probabilities, or restricted mean survival times, for different treatment choices. Flexibility to generate patient specific predictions are important in this setting to allow for personalized treatment selection. As a Bayesian approach, NFT BART offers posterior uncertainty summaries for the prediction inference, a benefit that may not be available with other machine learning approaches. We applied the proposed approach to the problem of optimal donor selection for a MUD HCT. Our approach can be readily extended to a general setting with a large number of patient specific treatment options. In the donor selection application patients have different sets of potential donors due to their different HLA typing, and some patients may have a very long list of potential donors to choose from. The main clinical conclusions of this study are that the youngest available male donor should generally be prioritized for all patients, except when there is a female donor considerably younger who would likely have better overall survival outcomes despite lower event-free survival. No other donor factors were important for either overall survival or event-free survival, acknowledging that there was lesser power for determining the impact of HLA-DQB1 due to limited numbers of mismatches on this locus. Although some patients may have many donors to choose from, the monotone effect of donor age and the reduced set of important donor characteristics simplified the optimal donor choice to just selecting between two donors (youngest male and youngest female). However, in general the framework that we used can be implemented even when the decision process does not simplify like this.

There are some limitations to this study. Our study focused on MUD transplants only, and the cohort had limited use of post-transplant cyclophosphamide (PTCy) as a GVHD prophylaxis strategy. PTCy is surging in popularity due to its successful use in matched and mismatched donor transplants. The main benefit of PTCy is in reduced acute and chronic GVHD leading to improvements in EFS, but with limited impact on OS. Future studies using our general strategy should expand to include mismatched donor transplants and consider the impact of PTCy in donor selection strategies. Additionally, there may be other donor factors to be considered in a selection algorithm; as data on these become available, their contribution to donor selection algorithms could be examined using our approach. Finally, we have focused here on an approach of weighting hierarchically ordered survival outcomes for optimization. Future work could investigate a multi-state prediction model with utilities for each state. This would help our understanding of the contribution of different donor characteristics to each of the components for the EFS endpoint.

## Data Availability

CIBMTR supports accessibility of research in accord with the National Institutes of Health (NIH) Data Sharing Policy and the National Cancer Institute (NCI) Cancer Moonshot Public Access and Data Sharing Policy. The CIBMTR only releases de-identified datasets that comply with all relevant global regulations regarding privacy and confidentiality.

https://cibmtr.org/CIBMTR/Resources/Publicly-Available-Datasets

## Acknowledgments

CIBMTR is supported primarily by the Public Health Service U24CA076518 from the National Cancer Institute (NCI), the National Heart, Lung and Blood Institute (NHLBI), and the National Institute of Allergy and Infectious Diseases (NIAID); 75R60222C00011 from the Health Resources and Services Administration (HRSA); and N00014-23-1-2057 and N00014-24-1-2057 from the Office of Naval Research. Support is also provided by the Medical College of Wisconsin, NMDP, Gateway for Cancer Research, Pediatric Transplantation and Cellular Therapy Consortium and from the following commercial entities: AbbVie; Actinium Pharmaceuticals, Inc.; Adaptive Biotechnologies Corporation; ADC Therapeutics; Adienne SA; Alexion; AlloVir, Inc.; Amgen, Inc.; Astellas Pharma US; AstraZeneca; Atara Biotherapeutics; BeiGene; BioLineRX; Blue Spark Technologies; bluebird bio, inc.; Blueprint Medicines; Bristol Myers Squibb Co.; CareDx Inc.; CSL Behring; CytoSen Therapeutics, Inc.; DKMS; Elevance Health; Eurofins Viracor, DBA Eurofins Transplant Diagnostics; Gamida-Cell, Ltd.; Gift of Life Biologics; Gift of Life Marrow Registry; GlaxoSmithKline; HistoGenetics; Incyte Corporation; Iovance; Janssen Research & Development, LLC; Janssen/Johnson & Johnson; Jasper Therapeutics; Jazz Pharmaceuticals, Inc.; Karius; Kashi Clinical Laboratories; Kiadis Pharma; Kite, a Gilead Company; Kyowa Kirin; Labcorp; Legend Biotech; Mallinckrodt Pharmaceuticals; Med Learning Group; Medac GmbH; Merck & Co.; Mesoblast; Millennium, the Takeda Oncology Co.; Miller Pharmacal Group, Inc.; Miltenyi Biotec, Inc.; MorphoSys; MSA-EDITLife; Neovii Pharmaceuticals AG; Novartis Pharmaceuticals Corporation; Omeros Corporation; OptumHealth; Orca Biosystems, Inc.; OriGen BioMedical; Ossium Health, Inc.; Pfizer, Inc.; Pharmacyclics, LLC, An AbbVie Company; PPD Development, LP; REGiMMUNE; Registry Partners; Rigel Pharmaceuticals; Sanofi; Sarah Cannon; Seagen Inc.; Sobi, Inc.; Stemcell Technologies; Stemline Technologies; STEMSOFT; Takeda Pharmaceuticals; Talaris Therapeutics; Vertex Pharmaceuticals; Vor Biopharma Inc.; Xenikos BV. The views expressed in this article do not reflect the official policy or position of the National Institute of Health, the Department of the Navy, the Department of Defense, Health Resources and Services Administration (HRSA) or any other agency of the U.S. Government.

