## Supplemental Information for "Optimal Donor Selection Across Multiple Outcomes For Hematopoietic Stem Cell Transplantation By Bayesian Nonparametric Machine Learning"

### 1 Introduction

For this study, we rely upon Nonparametric Failure Time Bayesian Additive Regression Trees (NFT BART) methodology (Sparapani et al., 2023). NFT BART is a nonparametric ensemble of trees survival analysis approach that does not rely on precarious restrictive assumptions such as proportionality, linearity nor homoskedasticity. These features are particularly pertinent for our study examining predictors for mortality risk in patients undergoing hematopoietic stem cell transplant (HSCT) for blood-borne cancer, where heteroskedasticity and non-proportional hazards are likely to be present.

Here, we give a general summary of NFT BART and its most relevant aspects for this study. For more details on NFT BART, see Sparapani et al. (2023) and the corresponding Web Supplement. In Section 2, we review the BART and Heteroskedastic BART (HBART) priors. Section 3 introduces the NFT BART model. Section 4 provides the default settings for the Low Information Omnibus Dirichlet Process Mixtures (LIO DPM) hierarchical prior. In Section 5, we provide DPM density, distribution, hazard and survival function estimates for NFT BART. In Section 6, we show how the restricted mean survival time (RMST) can be calculated for an NFT BART model. In Section 7, we demonstrate how to summarize an NFT BART model with marginal effects by Friedman's partial dependence (FPD) function. In Section 8, we provide a brief description of the **nftbart** R package. You will find additional details about model variables in Section 9.1 and their relationship of with OS/EFS in Section 9.2.

### 2 BART and Heteroskedastic BART

#### 2.1 The BART prior

BART (Chipman et al., 2010) is a sum of binary trees nonparametric machine learning regression model where the relationship between the outcome, $y_i$, and the covariates, $\boldsymbol{x}_i$, is learned from the data itself. We restrict our attention to a continuous outcomes for now. Let $y_i$ be a continuous outcome with $i = 1, \ldots, N$ indexing subjects and $\boldsymbol{x}_i$ is a vector of covariates. The BART model has the following form:

$$
\begin{aligned}
y_i \;=\; \mu(\boldsymbol{x}_i) + \epsilon_i \qquad & \mu \overset{\text{prior}}{\sim} \text{BART}(a, b, H, \kappa, \widetilde{\mu}) \qquad \mu(\boldsymbol{x}_i) \equiv \widetilde{\mu} + \sum_{h=1}^{H} g(\boldsymbol{x}_i; \mathcal{T}_h, \mathcal{L}_h) \\
\epsilon_i | \sigma^2 \overset{\text{iid}}{\sim} \text{N}\big(0,\ w_i^2 \sigma^2\big) \qquad & \sigma^2 \overset{\text{prior}}{\sim} \lambda \nu \chi^{-2}(\nu)
\end{aligned} \tag{1}
$$

where $\widetilde{\mu}$ is a constant that centers the data (a default choice is $\widetilde{\mu} = \overline{y}$). The binary tree regression function is $g(\boldsymbol{x}; \mathcal{T}, \mathcal{L})$ with $\mathcal{T}$ denoting the nodes of the tree (as well as whether they are branch decision rules or leaves) and $\mathcal{L}$ denoting its leaf values. The variance multiples, $w_i^2$, are known constants dictated by the problem of interest; when unneeded, simply let $w_i \equiv 1$. Prior argument default settings are employed that often provide adequate fits. For example, the number of trees, $H$, is *large* with typical settings of 50, 100 or 200 where 50 is a default with reasonable performance (Bleich et al., 2014).

#### 2.2 The Heteroskedastic BART (HBART) prior

Heteroskedastic BART (Pratola et al., 2020) is an extension to BART where we fit both a mean function, $\mu$, and a variance function, $\sigma^2$, for a flexible nonparametric function of $\boldsymbol{x}$. This model can be written as follows:

$$
\begin{aligned}
y_i \;=\; \mu(\boldsymbol{x}_i) + \epsilon_i \qquad & \mu \overset{\text{prior}}{\sim} \text{BART}(a, b, H, \kappa, \widetilde{\mu}) \qquad \mu(\boldsymbol{x}_i) \equiv \widetilde{\mu} + \sum_{h=1}^{H} g(\boldsymbol{x}_i; \mathcal{T}_h, \mathcal{L}_h) \\
\epsilon_i | \sigma^2 \overset{\text{ind}}{\sim} \text{N}\big(0,\ w_i^2 \sigma^2(\boldsymbol{x}_i)\big) \quad & \sigma^2 \overset{\text{prior}}{\sim} \text{HBART}(\widetilde{a}, \widetilde{b}, \widetilde{H}, \widetilde{\lambda}, \widetilde{\nu}) \quad \sigma^2(\boldsymbol{x}_i) \equiv \prod_{h=1}^{\widetilde{H}} g(\boldsymbol{x}_i; \widetilde{\mathcal{T}}_h, \widetilde{\mathcal{L}}_h)
\end{aligned} \tag{2}
$$

where the covariates of $\mu(.)$ and $\sigma^2(.)$ do not need to be the same, but we denote them by $\boldsymbol{x}_i$ merely for notational convenience throughout. Recommended prior default argument settings often provide adequate fitting: $\kappa = 5$ and $\widetilde{\mu} = \overline{y}$ for $\mu$; $\widetilde{\lambda} = s_y^2$ and $\widetilde{\nu} = 10$ for $\sigma^2$; with $a = \widetilde{a} = 0.95$ and $b = \widetilde{b} = 2$ for both.

#### 2.3 Binary tree regression models

Here we present a brief overview of binary tree regression models as well as delving into the details for the BART and HBART priors. $[\theta]$ is the *generic bracket notation* of Gelfand and Smith (1990) denoting the distribution of $\theta$, e.g., the prior for $\theta$ is $[\theta]$, the likelihood $[y|\theta]$ and the posterior $[\theta|y]$. For the BART prior, it is assumed that the trees are independent and the leaves are conditionally independent given the trees as $[\boldsymbol{\mathcal{T}}, \boldsymbol{\mathcal{L}}] \overset{\text{prior}}{\propto} \prod_h [\mathcal{T}_h] [\mathcal{L}_h | \mathcal{T}_h]$ where $\mathcal{T}_h$ represents a tree and $\mathcal{L}_h$ the leaves of that tree (the HBART prior is analogous: $\left[\widetilde{\boldsymbol{\mathcal{T}}}, \widetilde{\boldsymbol{\mathcal{L}}}\right]$).

#### 2.4 The variance prior for BART

The prior variance is independent of the trees and leaves. It is weakly informed by the data via the construction of $\widehat{\sigma}^2$ that defaults to either the variance of $y$ itself or the variance of the random error from a linear regression. A default value of $\nu = 3$ is assumed and $\lambda$ is chosen given the distribution, $\sigma^2 \overset{\text{prior}}{\sim} \lambda\nu\chi^{-2}(\nu)$, such that $\mathrm{P}[\sigma^2 < \widehat{\sigma}^2] = q$ where the default for $q = 0.9$. For more details of the variance prior see Chipman et al. (2010).

#### 2.5 The tree prior for BART and HBART

To develop the tree prior, we need to introduce some notation: $[\boldsymbol{\mathcal{T}}_h] = [\beta_{h1}, \ldots, \beta_{hC_h}]$ where $n = 1, \ldots, C_h$ is the index for nodes. For each node, the depth of the tree can be deduced from the node index itself: $d(n) = \lfloor \log_2 n \rfloor$. Furthermore, at tier $d(n) = d$, the potential nodes are numbered as $n = 2^d, \ldots, 2^{d+1} - 1$ from left to right where a parent's children are $2n$ and $2n + 1$ at depth $d + 1$ (but of course they need not all exist except $n = 1$). Rather than a tier's height, it is tier depth to be considered since binary trees are typically drawn *growing* downward (as opposed to upward like wooden trees); see the schematic diagram: Supplement Table 1.

| Tier | Node | | | | | | |
|---|---|---|---|---|---|---|---|
| 0 | | | | 1 | | | |
| 1 | | 2 | | | | 3 | |
| 2 | 4 | | 5 | | 6 | | 7 |
| ⋮ | | | | | | | |
| $d$ | | $2^d$ | | $\ldots$ | $2^{d+1}-1$ | | |

Supplement Table 1: Schematic diagram of a binary tree.

Trees with many branches will likely over-fit to the training data at the expense of predictive performance on unseen validation data without a regularization penalty. Therefore, the BART and HBART priors have a *branching penalty* where the regularity increases with the depth. If we assume a *success* is a branch decision rule and a *failure* is a terminal leaf value, then we have independent Bernoulli random variables $\beta_{hn} \overset{\text{prior}}{\sim} \mathrm{B}(p(d(n)))$ where $p(d) = a(1 + d)^{-b}$ and $q(d) = 1 - p(d)$. The prior argument defaults are $a = 0.95$ and $b = 2$ (for a discussion of this penalty, see Chipman et al. (1998, 2010); Ročková and Saha (2019)). So the expected number of branches (leaves), in prior probability, is 1 (2) with probability $\mathrm{P}[\beta_{h1} = 1, \beta_{h2} = \beta_{h3} = 0] = p(0)q(1)^2 \approx 0.55$ and 2 (3) with probability $2\mathrm{P}[\beta_{h1} = \beta_{h2} = 1, \beta_{h3} = \beta_{h4} = \beta_{h5} = 0] = 2p(0)p(1)q(1)q(2)^2 \approx 0.27$ (doubled due to symmetry), i.e., trees with only 1 or 2 branches (2 or 3 leaves) would predominate in prior probability of roughly 0.82.

#### 2.6 The leaf prior

##### 2.6.1 BART

A priori, each of the leaf values are independent: $[\mathcal{L}_h|\mathcal{T}_h] = \prod_{l_h} [\mu_{hl_h}|\mathcal{T}_h]$ where $\mathcal{L}_h = [\mu_{hl_h}]$ is a vector of leaf values for tree $h$ and $l_h \in \{n : \beta_{hn} = 0\}$ is the index for the leaves. Suppose that $y \in [y_{\min}, y_{\max}]$ and denote $\widetilde{\mu}_h$ as the leaf output values from each tree corresponding to the vector of covariates, $\boldsymbol{x}$. If $\widetilde{\mu}_h|\mathcal{T}_h \overset{\text{iid}}{\sim} \mathrm{N}\left(0,\ \sigma_\mu^2\right)$, then the model estimate is $\widehat{y} = \mathrm{E}\left[y\right] = \widetilde{\mu} + \sum_h \widetilde{\mu}_h$ where $\widehat{y} \sim \mathrm{N}\left(\widetilde{\mu},\ H\sigma_\mu^2\right)$. We choose a value for $\sigma_\mu$ which is the solution to the equations $y_{\min} = \widetilde{\mu} - \kappa\sqrt{H}\sigma_\mu$ and $y_{\max} = \widetilde{\mu} + \kappa\sqrt{H}\sigma_\mu$, i.e., $\sigma_\mu = \frac{y_{\max} - y_{\min}}{2\kappa\sqrt{H}}$. Therefore, we arrive at $\mu_{hl_h}|\mathcal{T}_h \overset{\text{prior}}{\sim} \mathrm{N}\left(0,\ \frac{\tau^2}{4\kappa^2 H}\right)$ where $\tau = y_{\max} - y_{\min}$. So, the default value, $\kappa = 2$, corresponds to $\widehat{y}$ falling within the extrema with approximately 0.95 probability. The values $y_{\min}$ and $y_{\max}$ can be elicited from expert opinion as the 2.5 and 97.5 percentiles respectively to set $\tau$. By default, $\tau$ is set to the extrema from the observed data.

##### 2.6.2 HBART

The BART leaf prior is as above except that the default is now $\kappa = 5$. Here, we discuss the prior for $\widetilde{\mathcal{L}}_h|\widetilde{\mathcal{T}}_h$. For a more detailed discussion of the HBART prior specification, please see Pratola et al. (2020). A priori, each of the leaf values are independent: $\left[\widetilde{\mathcal{L}}_h|\widetilde{\mathcal{T}}_h\right] = \prod_{l_h} \left[\sigma_{hl_h}^2|\widetilde{\mathcal{T}}_h\right]$ where $\widetilde{\mathcal{L}}_h = \left[\sigma_{hl_h}^2\right]$ is a vector of leaf values for tree $h$ and $l_h \in \{n : \widetilde{\beta}_{hn} = 0\}$ is the index for the leaves. The prior for the leaves is $\sigma_{hl_h}^2 \overset{\text{prior}}{\sim} \lambda\nu\chi^{-2}\left(\nu\right)$ so we need to determine the defaults for $\lambda$ and $\nu$. Suppose that we consider a homoskedastic BART model: $\mathrm{E}\left[\sigma^2\right] = \frac{\widetilde{\lambda}\widetilde{\nu}}{\widetilde{\nu}-2}$ with defaults $\widetilde{\lambda} = \widehat{\sigma}^2$ and $\widetilde{\nu} = 10$. Compare that with the corresponding HBART setting: $\mathrm{E}\left[\sigma^2(\boldsymbol{x})\right] = \prod_h \mathrm{E}\left[\widetilde{\sigma}_h^2\right] = \lambda^{\widetilde{H}}\left[\frac{\nu}{\nu-2}\right]^{\widetilde{H}}$ (where $\widetilde{\sigma}_h^2$ represents the leaf from tree $h$ corresponding to $\boldsymbol{x}$). Since $\frac{\widetilde{\lambda}\widetilde{\nu}}{\widetilde{\nu}-2} \equiv \lambda^{\widetilde{H}}\left[\frac{\nu}{\nu-2}\right]^{\widetilde{H}}$, we use the values $\lambda = \widetilde{\lambda}^{1/\widetilde{H}}$ and $\nu = 2\left[1 - \left(1 - \frac{2}{\widetilde{\nu}}\right)^{1/\widetilde{H}}\right]^{-1}$.

### 3 NFT BART

NFT BART is a largely assumption free survival analysis model having a nonparametric distribution for the random error while allowing the covariates to explain both a location shift and a scale change. Here we present a brief introduction; for more details see Sparapani et al. (2023). For notational convenience, we move fluidly between a parameterization based on the precision, $\tau_i$, to the variance, $\sigma_i^2 = \tau_i^{-1}$, since it is generally arbitrary. The NFT BART model is as follows (let the unknown parameters be denoted $\boldsymbol{\theta}$):

$$\begin{Bmatrix} y_i & = & \mu(\boldsymbol{x}_i) + \epsilon_i & \mu & \overset{\text{prior}}{\sim} & \mathrm{BART}(a, b, H, \kappa, \widetilde{\mu}) \\ \epsilon_i|\boldsymbol{\theta} & \overset{\text{ind}}{\sim} & \mathrm{N}(\mu_i,\ \sigma_i^2\sigma^2(\boldsymbol{x}_i)) & \sigma^2 & \overset{\text{prior}}{\sim} & \mathrm{HBART}(\widetilde{a}, \widetilde{b}, \widetilde{H}, \widetilde{\lambda}, \widetilde{\nu}) \end{Bmatrix} \tag{3}$$

subject to the constraints $N^{-1}\sum_i \mu_i = 0$ and $N^{-1}\sum_i \sigma_i^2 = 1$ for identifiability (we defer the description of priors for $\mu_i$ and $\tau_i$ until the next Section 4). Right-censoring is handled by data augmentation as follows:

$$y_i \begin{cases} \sim \mathrm{N}(\mu_i + \mu(\boldsymbol{x}_i),\ \sigma_i^2\sigma^2(\boldsymbol{x}_i))\,\mathrm{I}(\log t_i, \infty) & \text{if } \delta_i = 0, \text{ right-censoring} \\ = \ \log t_i & \text{if } \delta_i = 1, \text{ an event time} \end{cases} \tag{4}$$

and, conveniently, left-censoring can be handled similarly, but that is not shown for simplicity.

As noted, both $\mu_i$ and $\sigma_i$ are constrained. Posterior inference can reliably be performed in the unconstrained setting; however, convergence diagnostics are more challenging without constraints. Furthermore, in the unconstrained setting, our experience has been that MCMC mixing is inefficient making convergence diagnostics even more important at the same time that they are more difficult to ascertain. Therefore, we recommend constrained DPM for NFT BART (Yang et al., 2010).

### 4 The LIO DPM prior hierarchy

Like BART and HBART, the LIO DPM prior hierarchy has robust default arguments that should be sufficient in most circumstances. Due to the identifiability constraints on $(\mu_i, \tau_i)$, the defaults fall into what is known as the standardized setting (Shi et al., 2019). The DPM LIO prior hierarchy employed by NFT BART is as follows:

$$\begin{aligned} G|\alpha &\overset{\text{prior}}{\sim} \mathrm{DP}\left(\alpha,\ F_{(\mu_0,\tau_0|k_0,b_0)}\right) & (\mu_i,\tau_i)|G &\overset{\text{prior}}{\sim} G & & \\ \alpha &\overset{\text{prior}}{\sim} \mathrm{Gamma}\,(1,\ 0.1) & \tau_0|b_0 &\underset{F}{\overset{\text{prior}}{\sim}} \mathrm{Gamma}\,(1.5,\ b_0) & b_0 &\overset{\text{prior}}{\sim} \mathrm{Gamma}\,(2,\ 1) \\ & & \mu_0|(\tau_0,k_0) &\underset{F}{\overset{\text{prior}}{\sim}} \mathrm{N}\big(0,\ \tau_0^{-1}k_0^{-1}\big) & k_0 &\overset{\text{prior}}{\sim} \mathrm{Gamma}\,(3,\ 7.5) \end{aligned} \tag{5}$$

where the joint base distribution $[\mu_0, \tau_0|k_0, b_0]$ is drawn by the conditionals $[\tau_0|b_0][\mu_0|\tau_0, k_0]$.

### 5 Posterior inference for NFT BART

Notationally, let $\boldsymbol{\theta} = (\mu, \sigma^2, \boldsymbol{\mu}, \boldsymbol{\tau}, \alpha)$ represent the unknown parameters where $\boldsymbol{\theta}_m$ is the $m$th posterior draw. Our primary interest with respect to statistical inference here is the impact of the covariates on the time to an event. In particular, the survival function, $S(t|\boldsymbol{x}) = 1 - F(t|\boldsymbol{x})$, plays a central role along with the hazard function, $h(t|\boldsymbol{x}) = f(t|\boldsymbol{x})/S(t|\boldsymbol{x})$. The nonparametric estimation of survival and hazard is arrived at by aggregating over the DPM clusters (Escobar and West, 1995) to create estimates of the distribution, $F(t|\boldsymbol{x})$, and density, $f(t|\boldsymbol{x})$. For NFT BART, we arrive at the following (conditioning on $\boldsymbol{\theta}_m$ suppressed for notational convenience):

$$F_m(t|\boldsymbol{x}) = \int \Phi\left\{\frac{\log t - \mu_* - \mu_m(\boldsymbol{x})}{\sigma_*\sigma_m(\boldsymbol{x})}\right\} G_m(\mathrm{d}\mu_*, \mathrm{d}\sigma_*) \tag{6}$$

$$= \sum_{j=1}^{\infty} \omega_j \Phi\left\{\frac{\log t - \mu_j^* - \mu_m(\boldsymbol{x})}{\sigma_j^*\sigma_m(\boldsymbol{x})}\right\} \approx \sum_{j=1}^{K_m} \omega_{jm} \Phi\left\{\frac{\log t - \mu_{jm}^* - \mu_m(\boldsymbol{x})}{\sigma_{jm}^*\sigma_m(\boldsymbol{x})}\right\}$$

$$f_m(t|\boldsymbol{x}) = \int \frac{\phi\left\{\frac{\log t - \mu_* - \mu_m(\boldsymbol{x})}{\sigma_*\sigma_m(\boldsymbol{x})}\right\}}{t\sigma_*\sigma_m(\boldsymbol{x})} G_m(\mathrm{d}\mu_*, \mathrm{d}\sigma_*) \tag{7}$$

$$= \sum_{j=1}^{\infty} \frac{\omega_j \phi\left\{\frac{\log t - \mu_j^* - \mu_m(\boldsymbol{x})}{\sigma_j^*\sigma_m(\boldsymbol{x})}\right\}}{t\sigma_j^*\sigma_m(\boldsymbol{x})} \approx \sum_{j=1}^{K_m} \frac{\omega_{jm} \phi\left\{\frac{\log t - \mu_{jm}^* - \mu_m(\boldsymbol{x})}{\sigma_{jm}^*\sigma_m(\boldsymbol{x})}\right\}}{t\sigma_{jm}^*\sigma_m(\boldsymbol{x})}$$

where $\Phi(.)$ and $\phi(.)$ are the standard Normal distribution and density functions, respectively, while $m = 1, \ldots, M$ indexes draws from the posterior. In equations (6) and (7), we adopt the notation $(\mu_*, \sigma_*)$ for constrained posterior predictive draws from $G_m$ and $(\mu_j^*, \sigma_j^*)$ for those values that are shared by the $j$th atomic cluster. But, drawing from a constrained $G_m$ requires unconstrained draws needing centering/scaling. Therefore, we propose a reasonable compromise for computational convenience by substituting constrained $(\mu_{jm}^*, \sigma_{jm}^*)$ draws from the posterior for $(\mu_j^*, \sigma_j^*)$ that provides fairly effective estimation performance.

Now we can calculate our survival function estimate by the mean with respect to the posterior as $\widehat{S}(t|\boldsymbol{x}) = M^{-1}\sum_m S_m(t|\boldsymbol{x})$ (and the hazard function is estimated similarly). Further, we can create $1 - 2\pi$ level credible intervals via the $\pi$ and $1 - \pi$ quantiles of the posterior, $(\widehat{S}_\pi(t|\boldsymbol{x}), \widehat{S}_{1-\pi}(t|\boldsymbol{x}))$, such that $\widehat{S}_p(t|\boldsymbol{x}) = S_{m_p}(t|\boldsymbol{x})$ where $m_p$ is the posterior draw corresponding to the $p = \pi$, or $p = 1 - \pi$, quantile respectively.

### 6 Restricted mean survival time (RMST)

The restricted mean survival time (RMST) is an alternative measure that is fairly interpretable (Royston and Parmar, 2013; Pak et al., 2017; Kloecker et al., 2020). The mean survival time requires an infinite integral: $\int_0^\infty S(s|\boldsymbol{x})\mathrm{d}s$. However, this is impractical, i.e., an observation period can't be lengthened until every patient passes away. Therefore, RMST limits the observation period up until time $\tau$: $\mathrm{RMST}(\tau) = \int_0^\tau S(s|\boldsymbol{x})\mathrm{d}s$.

To calculate RMST from an NFT BART model, consider a log-Normal time-to-event, $\mathrm{e}^z$, where $z \sim \mathrm{N}(\mu,\ \sigma^2)$. This leads to the following intermediate result (a completing the square calculation reminiscent of the Moment Generating Function identity).

$$\begin{aligned}
\mathrm{E}\left[\mathrm{e}^{z}|z<\log(\tau)\right] &= (\sigma\sqrt{2\pi})^{-1}\int_{-\infty}^{\log(\tau)} \mathrm{e}^{z-\frac{(z-\mu)^2}{2\sigma^2}}\,dz \\
&= \mathrm{e}^{\mu+\sigma^2/2}(\sigma\sqrt{2\pi})^{-1}\int_{-\infty}^{\log(\tau)} \mathrm{e}^{-\frac{(z-(\mu+\sigma^2))^2}{2\sigma^2}}\,dz \\
&= \mathrm{e}^{\mu+\sigma^2/2}\Phi\left(\frac{\log(\tau)-(\mu+\sigma^2)}{\sigma}\right)
\end{aligned}$$

For NFT BART, we have a DP mixture of log-Normals: $\log(t) = z^*$ where $z^* \sim \sum_j \omega_j \,[z_j]$ and $[z_j]$ is $\mathrm{N}\big(\mu_j,\ \sigma_j^2\big)$. Now, we need the probability, $p$, that $z^*$ falls within the observation period.

$$p = \mathrm{P}[z^* < \log(\tau)] = \sum_j \omega_j \int_{-\infty}^{\log(\tau)} [z_j]\,\mathrm{d}z_j = \sum_j \omega_j p_j \text{ where } p_j = \Phi\left[\frac{\log(\tau)-\mu_j}{\sigma_j}\right]$$

We derive this next key result by recognizing that the RMST is simply the expectation of the following random variable: $s = \min(t, \tau)$.

$$\begin{aligned}
\mathrm{RMST}(\tau) &= \mathrm{E}\left[s\right] \\
&= \int_{-\infty}^{\log(\tau)} \mathrm{e}^{z^*}\,[z^*]\,\mathrm{d}z^* + \tau\int_{\log(\tau)}^{\infty} [z^*]\,\mathrm{d}z^* \\
&= \mathrm{E}\left[\mathrm{e}^{z^*}|z^*<\log(\tau)\right] + q\tau \text{ where } q = 1-p
\end{aligned}$$

This result can be decomposed into its atoms step-by-step.

$$\begin{aligned}
\mathrm{E}\left[\mathrm{e}^{z^*}|z^*<\log(\tau)\right] &= \sum_j \omega_j \mathrm{E}\left[\mathrm{e}^{z_j}|z_j<\log(\tau)\right] \\
\mathrm{E}\left[\mathrm{e}^{z_j}|z_j<\log(\tau)\right] &= \pi_j \mathrm{e}^{\mu_j+\sigma_j^2/2} \text{ where } \pi_j = \Phi\left(\frac{\log(\tau)-(\mu_j+\sigma_j^2)}{\sigma_j}\right) \\
\mathrm{E}\left[\mathrm{e}^{z^*}|z^*<\log(\tau)\right] &= \sum_j \omega_j \pi_j \mathrm{e}^{\mu_j+\sigma_j^2/2}
\end{aligned}$$

So, finally, arriving at the result of interest: $\mathrm{RMST}(\tau) = \mathrm{E}\left[s\right] = q\tau + \sum_j \omega_j \pi_j \mathrm{e}^{\mu_j+\sigma_j^2/2}$.

### 7 Marginal effects

Friedman's partial dependence function (FPD) is a common choice for estimating marginal effects via nonparametric regression and/or machine learning applications (Friedman, 2001). We divide the covariates into a subset of interest, $A$, and their complement, $B$, where all covariates are $A \cup B$. The covariates of interest are fixed at settings of interest: a single setting denoted $\boldsymbol{x}_A$. The complement take on the observed values found in the training data set, denoted $\boldsymbol{x}_{iB}$ for subject $i$, with the corresponding setting for all covariates denoted as $(\boldsymbol{x}_A, \boldsymbol{x}_{iB})$.

Notationally, let the unknown parameters be $\boldsymbol{\theta} = (\mu, \sigma^2, \boldsymbol{\mu}, \boldsymbol{\tau}, \alpha)$ where $\boldsymbol{\theta}_m$ is for the $m$th posterior draw; and, similarly, let $\boldsymbol{\theta}_i = (\mu, \sigma^2, \mu_i, \tau_i, \alpha)$ with corresponding $\boldsymbol{\theta}_{im}$. Formally, we have an interest in the marginal expectation $\widehat{y}_A = \mathrm{E}\left[y|\boldsymbol{x}_A\right]$. So, consider the draws of $\widehat{y}_{Am} = \int \mathrm{E}\left[y|\boldsymbol{x}_A, \boldsymbol{x}_B, \boldsymbol{\theta}_m\right] \left[\boldsymbol{x}_B|\boldsymbol{x}_A\right] \mathrm{d}\boldsymbol{x}_B$. Full evaluation of this conditional expectation can be challenging in practice due to the need for characterizing the conditional distribution $[\boldsymbol{x}_B|\boldsymbol{x}_A]$. The FPD function approximates this quantity under an assumption of independence by averaging over the observed marginal covariate distribution of $\boldsymbol{x}_B$ to get $\widehat{y}_{Am} \approx N^{-1} \sum_i \mathrm{E}\left[y|\boldsymbol{x}_A, \boldsymbol{x}_{iB}, \boldsymbol{\theta}_m\right]$ for the $m$th posterior draw. We conceptually extend the FPD technique to an NFT BART marginal expectation by the following simple adaptation: $\widehat{y}_{Am} \approx N^{-1} \sum_i \mathrm{E}\left[y_i|\boldsymbol{x}_A, \boldsymbol{x}_{iB}, \boldsymbol{\theta}_{im}\right] = N^{-1} \sum_i \left(\mu_{im} + \mu_m(\boldsymbol{x}_A, \boldsymbol{x}_{iB})\right)$ leading to the result $\widehat{y}_A = M^{-1} \sum_m \widehat{y}_{Am}$. Similarly, FPD can be employed for more complex functions of the posterior such as the survival. We arrive at the survival marginal effect for setting $\boldsymbol{x}_A$ for NFT BART, in two steps, as follows (conditioning on $\boldsymbol{\theta}$ suppressed for notational convenience).

$$\left\{\begin{array}{ll} F_{Am}(t|\boldsymbol{x}_A) & = N^{-1} \sum_i \Phi\left(\frac{\log t - \mu_{im} - \mu_m(\boldsymbol{x}_A, \boldsymbol{x}_{iB})}{\sigma_{im}\sigma_m(\boldsymbol{x}_A, \boldsymbol{x}_{iB})}\right) \\ \widehat{S}_A(t|\boldsymbol{x}_A) & = 1 - M^{-1} \sum_m F_{Am}(t|\boldsymbol{x}_A) \end{array}\right\}$$

And, finally, credible intervals for the marginal effects are provided by the posterior quantiles as shown above.

### 8 Software Implementation

The software necessary to implement the methodology explored in this article is not trivial to implement. We relied upon the **nftbart** R package that is publicly available online hosted on the Comprehensive R Archive Network (CRAN) (Sparapani et al., 2023). The **nftbart** package relied on several key computational methods some of which were explored in this article. The next section demonstrates an example discussing missing data imputation and the marginal effects methodology employed here. Furthermore, the Gibbs conditionals necessary for NFT BART are shown in the previous section. Other computational methods employed, besides BART (Chipman et al., 2010) and HBART (Pratola et al., 2020), include efficient BART/HBART posterior sampling (Pratola, 2016), efficient DPM sampling (Neal, 2000), constrained DPM (Yang et al., 2010), DPM LIO (Shi et al., 2019) and data augmentation for left-/right-censoring (Henderson et al., 2020).

With the **nftbart** R package, we present a real data example of an advanced lung cancer study (Loprinzi et al., 1994). Two-hundred and twenty-eight subjects with lung cancer were followed by the North Central Cancer Treatment Group for a median of roughly one year. Several covariates of interest were collected including age, sex, daily activity performance scores, diet and weight-loss information. All of these variables were largely non-missing with the exception of the calories consumed at meals for which missingness was 20.6%.

For this limited amount of missing data, we utilized record-level *cold-decking imputation* that is biased towards the null. The name reflects its similarity to hot-decking (de Waal et al., 2011) except that no attempt is made to locate a nearby/hot neighbor based on the outcome nor any other covariate criteria, i.e., cold-decking is a simple random sampling of a non-missing subject's record to replace the missing values with. For subject's with multiple missing values, the joint relationships between covariates are maintained by replacing all of the missing values from the non-missing subject randomly chosen. This simple missing data imputation method is sufficient for data sets with relatively few missing values; for more prevalent missingness we recommend the *sequential* BART algorithm (Xu et al., 2016).

For this example, sex was determined to be the most important covariate by Thompson Sampling Variable Selection (TSVS) (Liu and Ročková, 2023; Sparapani et al., 2023) with 138 male and 90 female participants. To demonstrate a common computation with **nftbart**, we will compare the survival experience of males vs. females by their marginal effects with Friedman's partial dependence function (Friedman, 2001) (as described in 7). As we can see in Supplement Figure 1, females generally have longer survival; however, for advanced lung cancer the prognosis is dire in the era of the collected data since the survival probability declines precipitously for both sexes. This demonstration is included with the **nftbart** package. You can install the **nftbart** R package and run this example as follows (use a nearby CRAN mirror for best results; see `http://cran.r-project.org/mirrors.html`).

```
> options(repos=c(CRAN="http://cran.r-project.org"))
> install.packages("nftbart", dependencies=TRUE)
> ## system.file() returns the location where lung.R is installed
> system.file("demo/lung.R", package="nftbart")
> source(system.file("demo/lung.R", package="nftbart"))
> ## demo("lung", package="nftbart") ## via the demo() facility
```

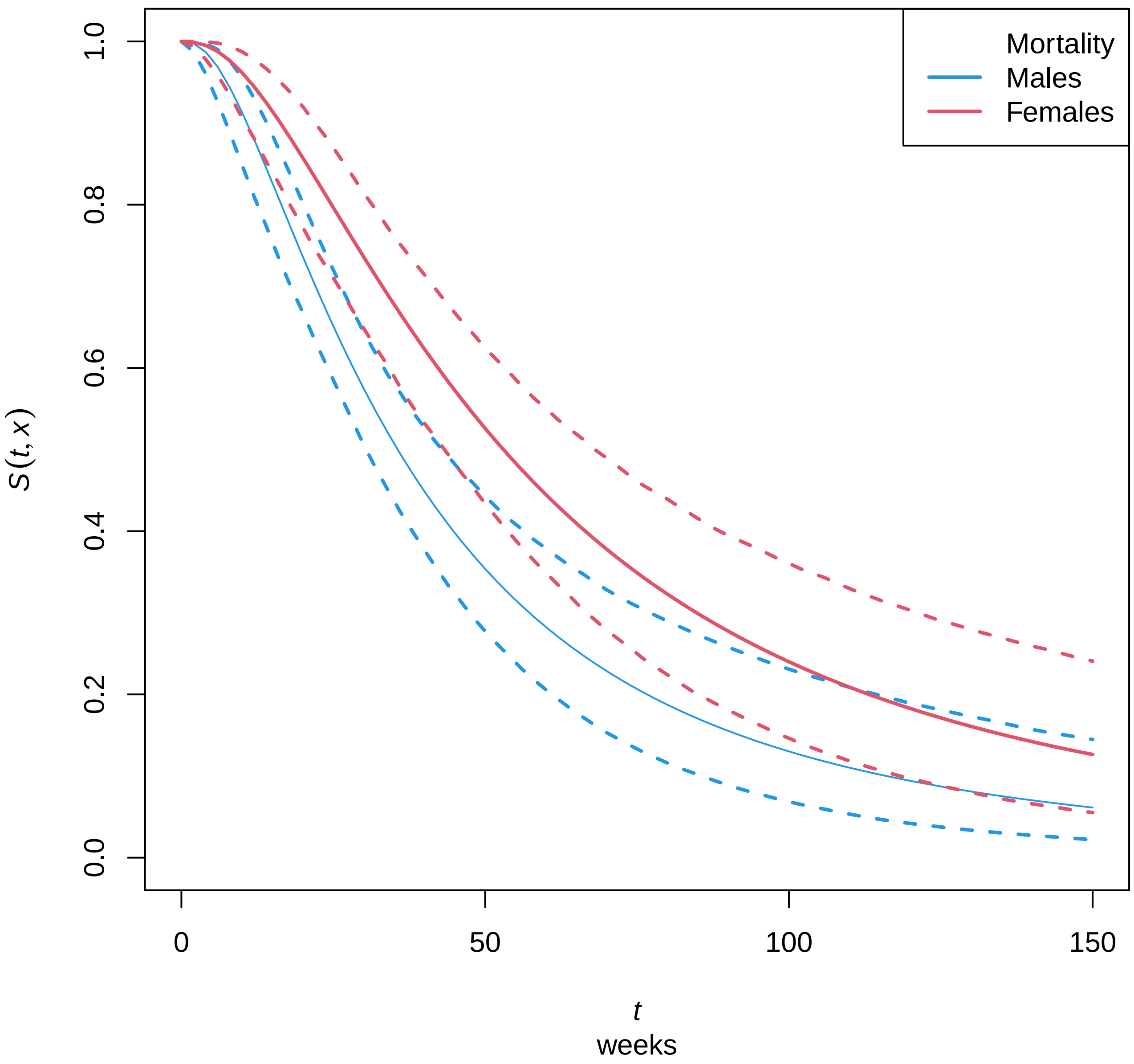


Supplement Figure 1: Advanced lung cancer study example: males vs. females. Two-hundred and twenty-eight subjects with lung cancer were followed by the North Central Cancer Treatment Group for a median of roughly one year: 138 male and 90 female participants. For this data set, statistical inference was performed with NFT BART for the collected covariates including age, sex, daily activity performance scores, diet and weight-loss information. The solid lines summarize the survival marginal effect for males (blue) and females (red) where the dashed lines are 95% credible intervals.

### 9 Additional details

#### 9.1 Model variables

First, we provide tabulations for the additional covariates in the model, 41 in total: Supplementary Tables 2, 3, 4, and 5. Furthermore, we have a comparison of the actual matched donor from the search archive along with two potential matches: the youngest female and the youngest male: Supplementary Tables 6.

This is a good point to elaborate on when cold-decking missing imputation was employed (as described in the previous section). For a global variable with missigness such as "Recipient CMV+", cold-decking was performed. And the same imputation per recipient was used for both OS and EFS calculations. However, for a disease-specific covariate like "ALL immunotype", this wouldn't work well. In this case, cold-decking would very likely just draw another missing value since only 13% of the recipients were transplanted due to ALL: we could keep drawing, but for a large data set (as we have here) that is time-consuming. So, for disease-specific variables such as these, we employed a missing value category rather than imputation.

Supplement Table 2: Recipient demographic characteristics. Training and validation sets are mutually exclusive while search archive is a subset of validation.

| | Training set | | Validation set | | Search archive | | Overall total | |
|---|---|---|---|---|---|---|---|---|
| **Recipient CMV+** | 9968 | | 1787 | | 691 | | 11755 | |
| Yes | 6021 | 60.4% | 1078 | 60.3% | 420 | 60.8% | 7099 | 60.4% |
| Missing | 48 | | 15 | | 8 | | 63 | |
| **Ventilation history** | 10012 | | 1801 | | 699 | | 11813 | |
| Yes | 367 | 3.7% | 67 | 3.7% | 23 | 3.3% | 434 | 3.7% |
| Missing | 4 | | 1 | | 0 | | 5 | |
| **Invasive fungal history** | 10009 | | 1801 | | 699 | | 11810 | |
| Yes | 421 | 4.2% | 83 | 4.6% | 40 | 5.7% | 504 | 4.3% |
| Missing | 7 | | 1 | | 0 | | 8 | |

CMV: cytomegalovirus

Supplement Table 3: Disease and transplant characteristics: part 1. Training and validation sets are mutually exclusive while search archive is a subset of validation.

| | Training set | | Validation set | | Search archive | | Overall total | |
|---|---|---|---|---|---|---|---|---|
| **Disease** | 10016 | | 1802 | | 699 | | 11818 | |
| ALL | 1290 | 12.9% | 251 | 13.9% | 102 | 14.6% | 1541 | 13.0% |
| AML | 3956 | 39.5% | 746 | 41.4% | 290 | 41.5% | 4702 | 39.8% |
| CLL | 139 | 1.4% | 26 | 1.4% | 10 | 1.4% | 165 | 1.4% |
| CML | 269 | 2.7% | 47 | 2.6% | 22 | 3.1% | 316 | 2.7% |
| MDS | 1988 | 19.8% | 362 | 20.1% | 139 | 19.9% | 2350 | 19.9% |
| MM | 128 | 1.3% | 26 | 1.4% | 10 | 1.4% | 154 | 1.3% |
| NHL | 617 | 6.2% | 94 | 5.2% | 37 | 5.3% | 711 | 6.0% |
| Other | 1629 | 16.3% | 250 | 13.9% | 89 | 12.7% | 1879 | 15.9% |
| **From diagnosis to transplant** | 9995 | | 1796 | | 695 | | 11791 | |
| 0:6 months | 4311 | 43.1% | 754 | 42.0% | 285 | 41.0% | 5065 | 43.0% |
| >6:12 months | 2511 | 25.1% | 475 | 26.4% | 186 | 26.8% | 2986 | 25.3% |
| >12:18 months | 785 | 7.9% | 151 | 8.4% | 59 | 8.5% | 936 | 7.9% |
| >18:24 months | 457 | 4.6% | 89 | 5.0% | 40 | 5.8% | 546 | 4.6% |
| >24 months | 1931 | 19.3% | 327 | 18.2% | 125 | 18.0% | 2258 | 19.2% |
| Missing | 21 | | 6 | | 4 | | 27 | |
| **Prior autologous** | 10016 | | 1802 | | 699 | | 11818 | |
| Yes | 480 | 4.8% | 87 | 4.8% | 44 | 6.3% | 567 | 4.8% |
| **ATG/Campath** | 10016 | | 1802 | | 699 | | 11818 | |
| Yes | 3855 | 38.5% | 696 | 38.6% | 296 | 42.3% | 4551 | 38.5% |
| **ALL/AML cycles to CR1** | 5246 | | 997 | | 392 | | 6243 | |
| 1 | 3607 | 68.8% | 674 | 67.6% | 274 | 69.9% | 4281 | 68.6% |
| 2 | 1044 | 19.9% | 225 | 22.6% | 86 | 21.9% | 1269 | 20.3% |
| 3+ | 595 | 11.3% | 98 | 9.8% | 32 | 8.2% | 693 | 11.1% |
| **ALL Ph chromosome** | 1290 | | 251 | | 102 | | 1541 | |
| Yes | 423 | 32.8% | 80 | 31.9% | 35 | 34.3% | 503 | 32.6% |
| **ALL immunotype** | 1279 | | 250 | | 101 | | 1529 | |
| T-cell | 181 | 14.2% | 37 | 14.8% | 16 | 15.8% | 218 | 14.3% |
| B-cell | 1098 | 85.8% | 213 | 85.2% | 85 | 84.2% | 1311 | 85.7% |
| Missing | 11 | | 1 | | 1 | | 12 | |
| **ALL cytogene** | 1290 | | 251 | | 102 | | 1541 | |
| Normal | 238 | 18.4% | 59 | 23.5% | 26 | 25.5% | 297 | 19.3% |
| Poor | 787 | 61.0% | 136 | 54.2% | 47 | 46.1% | 923 | 59.9% |
| Other | 265 | 20.5% | 56 | 22.3% | 29 | 28.4% | 321 | 20.8% |
| **ALL BCR/ABL marker** | 1290 | | 251 | | 102 | | 1541 | |
| Yes | 207 | 16.0% | 44 | 17.5% | 21 | 20.6% | 251 | 16.3% |

ABL: Abelson interactor 1 gene, ALL: acute lymphoblastic leukemia,
AML: acute myelogenous leukemia, ATG: anti-thymocyte globulin,
BCR: breakpoint cluster region gene, CLL: chronic lymphocytic leukemia,
CML: chronic myelogenous leukemia, CR1: first complete remission,
MDS: myelodysplastic syndrome, MM: multiple myeloma, NHL: non-Hodgkin's lymphoma.

Supplement Table 4: Disease and transplant characteristics: part 2. Training and validation sets are mutually exclusive while search archive is a subset of validation.

| | Training set | | Validation set | | Search archive | | Overall total | |
|---|---|---|---|---|---|---|---|---|
| **AML cytogene** | 3956 | | 746 | | 290 | | 4702 | |
| Normal | 259 | 6.5% | 60 | 8.0% | 31 | 10.7% | 319 | 6.8% |
| Favorable | 789 | 19.9% | 133 | 17.8% | 47 | 16.2% | 922 | 19.6% |
| Intermediate | 1249 | 31.6% | 254 | 34.0% | 102 | 35.2% | 1503 | 32.0% |
| Poor | 1641 | 41.5% | 293 | 39.3% | 107 | 36.9% | 1934 | 41.1% |
| APL | 18 | 0.5% | 6 | 0.8% | 3 | 1.0% | 24 | 0.5% |
| **AML progression from MDS** | 3956 | | 746 | | 290 | | 4702 | |
| Yes | 550 | 13.9% | 101 | 13.5% | 43 | 14.8% | 651 | 13.8% |
| **AML therapy-related** | 3956 | | 746 | | 290 | | 4702 | |
| Yes | 306 | 7.7% | 66 | 8.8% | 20 | 6.9% | 372 | 7.9% |
| **MDS predisposed** | 1988 | | 362 | | 139 | | 2350 | |
| Yes | 99 | 5.0% | 11 | 3.0% | 6 | 4.3% | 110 | 4.7% |
| **MDS therapy-related** | 1988 | | 362 | | 139 | | 2350 | |
| Yes | 385 | 19.4% | 57 | 15.7% | 23 | 16.5% | 442 | 18.8% |

AML: acute myelogenous leukemia, APL: acute promyelocytic leukemia,
MDS: myelodysplastic syndrome.

Supplement Table 5: Disease and transplant characteristics: part 3. Training and validation sets are mutually exclusive while search archive is a subset of validation.

| | Training set | | Validation set | | Search archive | | Overall total | |
|---|---|---|---|---|---|---|---|---|
| **CLL status** | 139 | | 26 | | 10 | | 165 | |
| Complete remission | 66 | 47.5% | 12 | 46.2% | 4 | 40.0% | 78 | 47.3% |
| Partial remission | 45 | 32.4% | 11 | 42.3% | 5 | 50.0% | 56 | 33.9% |
| Stable | 28 | 20.1% | 3 | 11.5% | 1 | 10.0% | 31 | 18.8% |
| **CLL 17p** | 139 | | 26 | | 10 | | 165 | |
| Yes | 50 | 36.0% | 11 | 42.3% | 4 | 40.0% | 61 | 37.0% |
| **CML status** | 269 | | 47 | | 22 | | 316 | |
| Hematologic CR | 99 | 36.8% | 17 | 36.2% | 7 | 31.8% | 116 | 36.7% |
| Chronic phase | 124 | 46.1% | 27 | 57.4% | 13 | 59.1% | 151 | 47.8% |
| Accelerated phase | 46 | 17.1% | 3 | 6.4% | 2 | 9.1% | 49 | 15.5% |
| **MM ISS/DS stage** | 128 | | 26 | | 10 | | 154 | |
| I/II | 45 | 35.2% | 11 | 42.3% | 6 | 60.0% | 56 | 36.4% |
| III | 83 | 64.8% | 15 | 57.7% | 4 | 40.0% | 98 | 63.6% |
| **MM cytorisk** | 103 | | 20 | | 7 | | 123 | |
| Normal | 7 | 6.8% | 3 | 15.0% | 0 | 0.0% | 10 | 8.1% |
| High | 70 | 68.0% | 8 | 40.0% | 5 | 71.4% | 78 | 63.4% |
| Standard | 26 | 25.2% | 9 | 45.0% | 2 | 28.6% | 35 | 28.5% |
| Missing | 25 | | 6 | | 3 | | 31 | |
| **MM status** | 128 | | 26 | | 10 | | 154 | |
| SCR/CR | 13 | 10.2% | 7 | 26.9% | 3 | 30.0% | 20 | 13.0% |
| VGPR | 63 | 49.2% | 11 | 42.3% | 4 | 40.0% | 74 | 48.1% |
| Partial response | 34 | 26.6% | 4 | 15.4% | 0 | 0.0% | 38 | 24.7% |
| Stable | 8 | 6.3% | 2 | 7.7% | 2 | 20.0% | 10 | 6.5% |
| Progressive/relapse | 10 | 7.8% | 2 | 7.7% | 1 | 10.0% | 12 | 7.8% |
| **NHL subtype** | 617 | | 94 | | 37 | | 711 | |
| Folicular | 80 | 13.0% | 7 | 7.4% | 2 | 5.4% | 87 | 12.2% |
| DLBCL | 188 | 30.5% | 42 | 44.7% | 17 | 45.9% | 230 | 32.3% |
| MCL | 95 | 15.4% | 11 | 11.7% | 3 | 8.1% | 106 | 14.9% |
| Other B-cell | 7 | 1.1% | 0 | 0.0% | 0 | 0.0% | 7 | 1.0% |
| T-cell | 247 | 40.0% | 34 | 36.2% | 15 | 40.5% | 281 | 39.5% |

CLL: chronic lymphocytic leukemia, CML: chronic myelogenous leukemia,
CR: complete response, DLBCL: diffuse large B-cell lymphoma,
ISS/DS: International Staging System/Durie-Salmon, (Hari et al., 2009)
MCL: mantle cell lymphoma, MM: multiple myeloma, NHL: non-Hodgkin's lymphoma,
SCR: stringent complete response, VGPR: very good partial response.

Supplement Table 6: Donor matching characteristics. Search archive comparison between the actual matched donor vs. potential matches: the youngest female, the youngest male and the youngest between the two.

| | Actual donor | | Youngest F | | Youngest M | | Youngest | |
|---|---|---|---|---|---|---|---|---|
| **Donor age** | 698 | | 699 | | 699 | | 699 | |
| 17:29 | 479 | 68.6% | 640 | 91.6% | 602 | 86.1% | 671 | 96.0% |
| 30:39 | 146 | 20.9% | 36 | 5.2% | 51 | 7.3% | 22 | 3.1% |
| 40:49 | 52 | 7.4% | 16 | 2.3% | 23 | 3.3% | 5 | 0.7% |
| 50:62 | 21 | 3.0% | 7 | 1.0% | 23 | 3.3% | 1 | 0.1% |
| Missing | 1 | 0 | | 0 | | | | |
| **Donor sex** | 698 | | 699 | | 699 | | 699 | |
| male | 524 | 75.1% | 0 | 0.0% | 699 | 100.0% | 347 | 49.6% |
| Missing | 1 | | 0 | | 0 | | 0 | |

#### 9.2 Relationships with respect to OS/EFS

Here we provide a table that cross-references each relationship with their corresponding data display(s).

Supplement Table 7: List of figures by assessed relationship.

| | OS | | EFS | |
|---|---|---|---|---|
| Characteristics | Marginal Effects | Waterfall Plot | Marginal Effects | Waterfall Plot |
| RMST value function | | | | |
| Figure 2 | | | | |
| Donor | | | | |
| Age | Figure 3 | | Figure 4 | |
| Sex/parity | Figure 5 | | Figure 6 | |
| Recipient female | | Figure 7 | | Figure 8 |
| Recipient male | | Figure 9 | | Figure 10 |
| CMV | Figure 11 | Figure 12 | Figure 13 | Figure 14 |
| DPB1 | Figure 15 | Figure 16 | Figure 17 | Figure 18 |
| DQB1 | Figure 19 | Figure 20 | Figure 21 | Figure 22 |
| Recipient | | | | |
| Age | Figure 23 | | Figure 24 | |
| Race | Figure 25 | | Figure 26 | |
| Median income (ZCTA) | Figure 27 | | Figure 28 | |
| HCT-CI | Figure 29 | | Figure 30 | |
| Disease/transplant | | | | |
| ALL status | Figure 31 | | Figure 32 | |
| AML status | Figure 33 | | Figure 34 | |
| MDS status | Figure 35 | | Figure 36 | |
| Year of transplant | Figure 37 | | Figure 38 | |

ALL: acute lymphoblastic leukemia, AML: acute myelogenous leukemia, CMV: cytomegalovirus,
HCT-CI: hematopoietic cell transplant comorbidity index (Sorror et al., 2015),
MDS: myelodysplastic syndrome, ZCTA: ZIP code tabulation area.

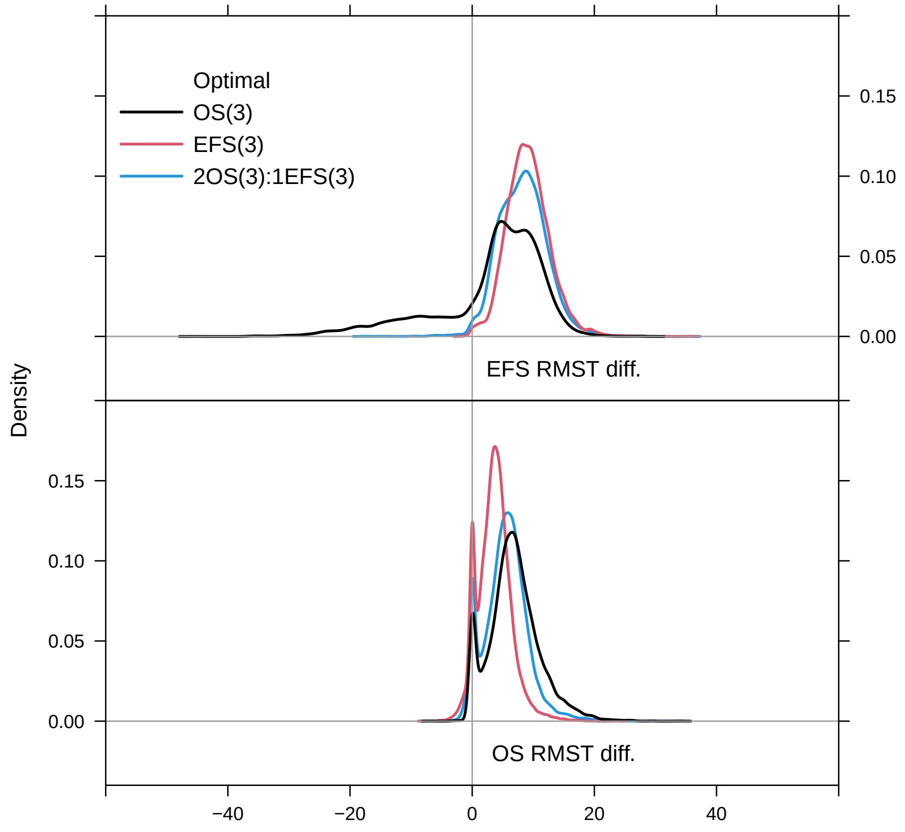


Supplement Figure 2: Population level value function for RMST. Return to cross-reference 9.2.

##### 9.2.1 Figures for donor characteristics

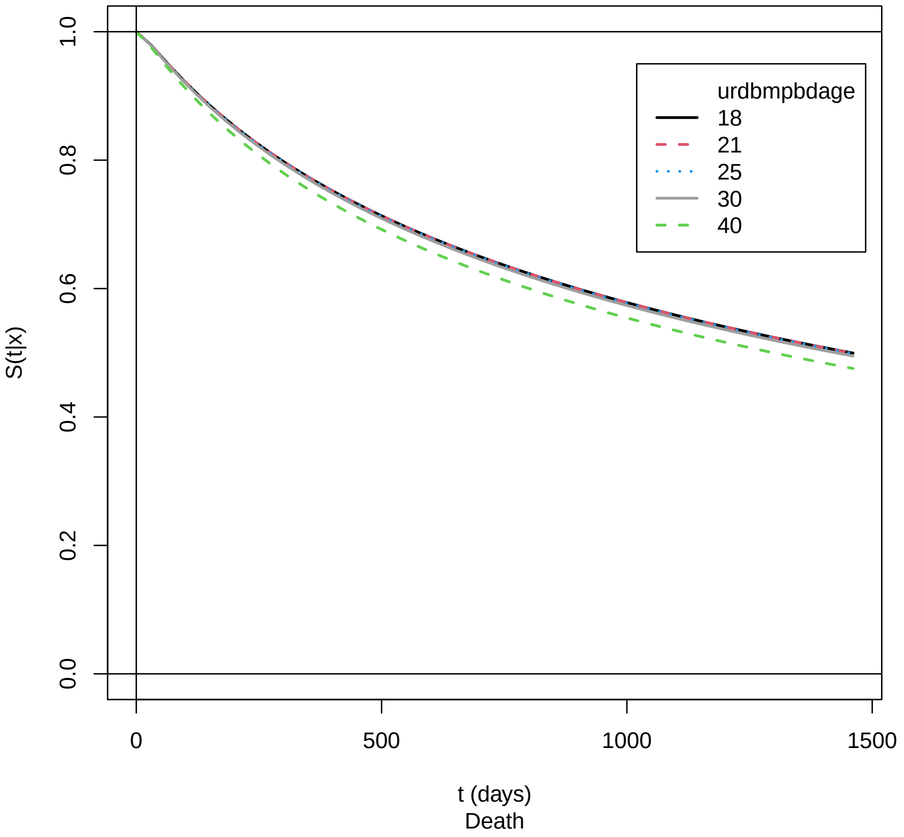


Supplement Figure 3: Marginal effect of donor age for OS: training cohort. Return to cross-reference 9.2.

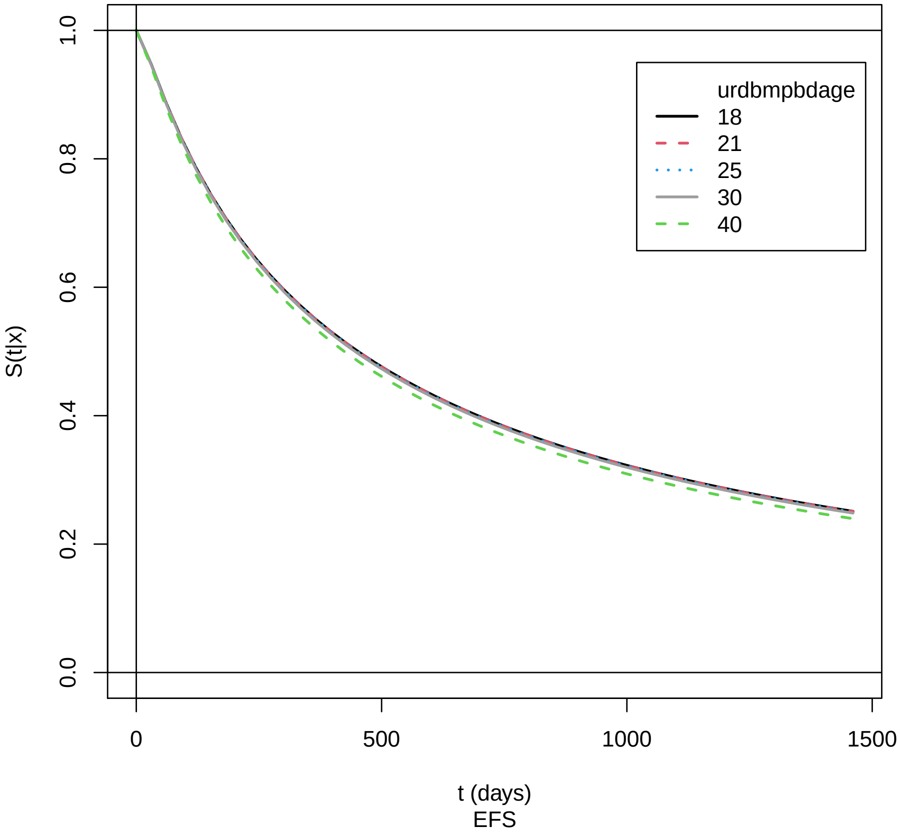


Supplement Figure 4: Marginal effect of donor age for EFS: training cohort. Return to cross-reference 9.2.

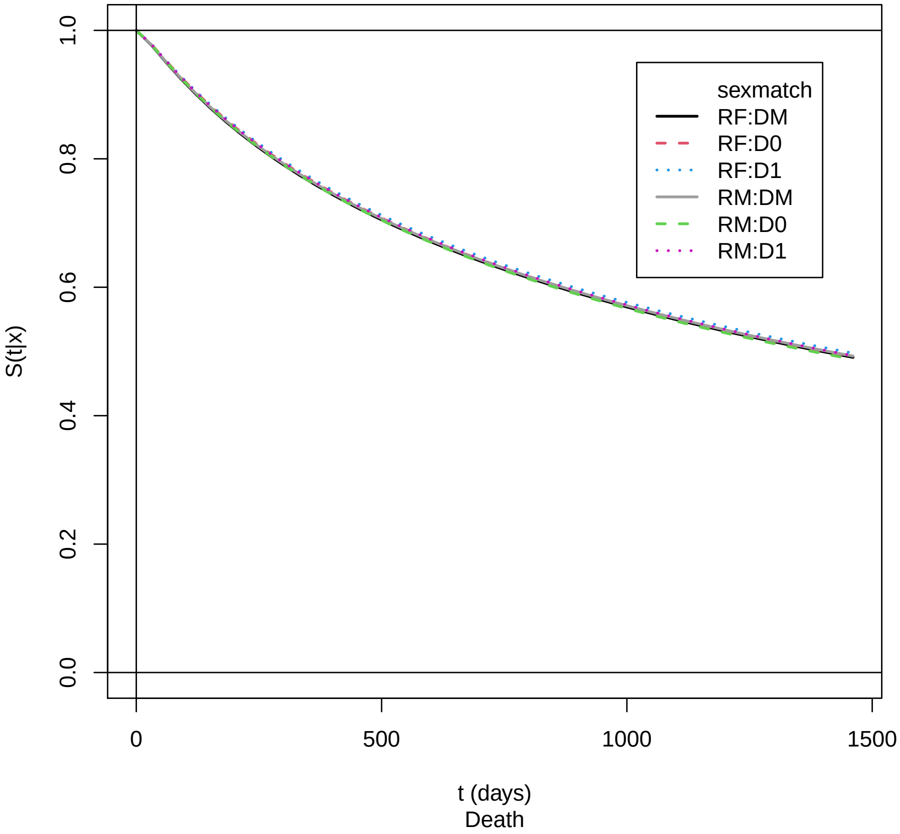


Supplement Figure 5: Marginal effect of sex match for OS: training cohort. Return to cross-reference 9.2.

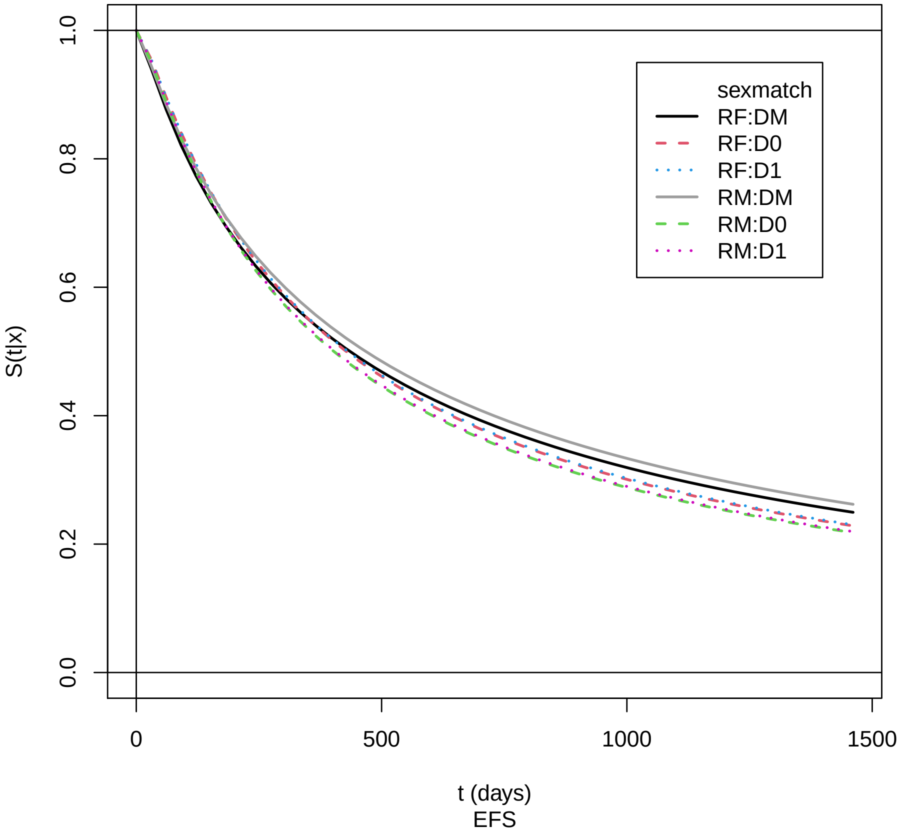


Supplement Figure 6: Marginal effect of sex match for EFS: training cohort. Return to cross-reference 9.2.

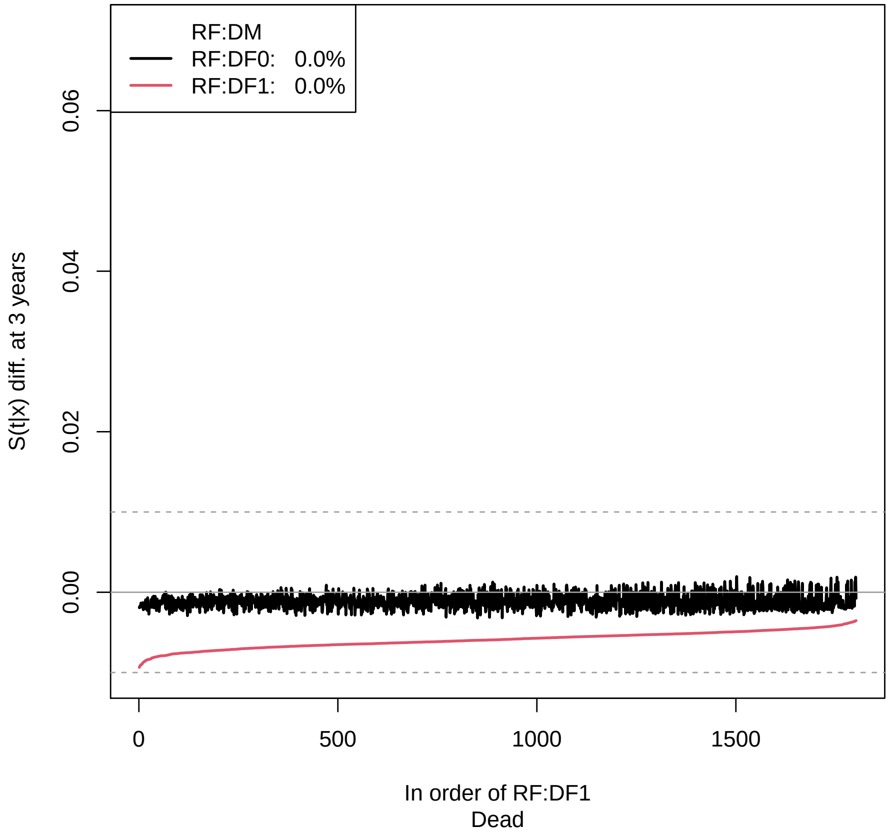


Supplement Figure 7: Waterfall plot of recipient females and donor sex/parity for OS: validation cohort. Return to cross-reference 9.2.

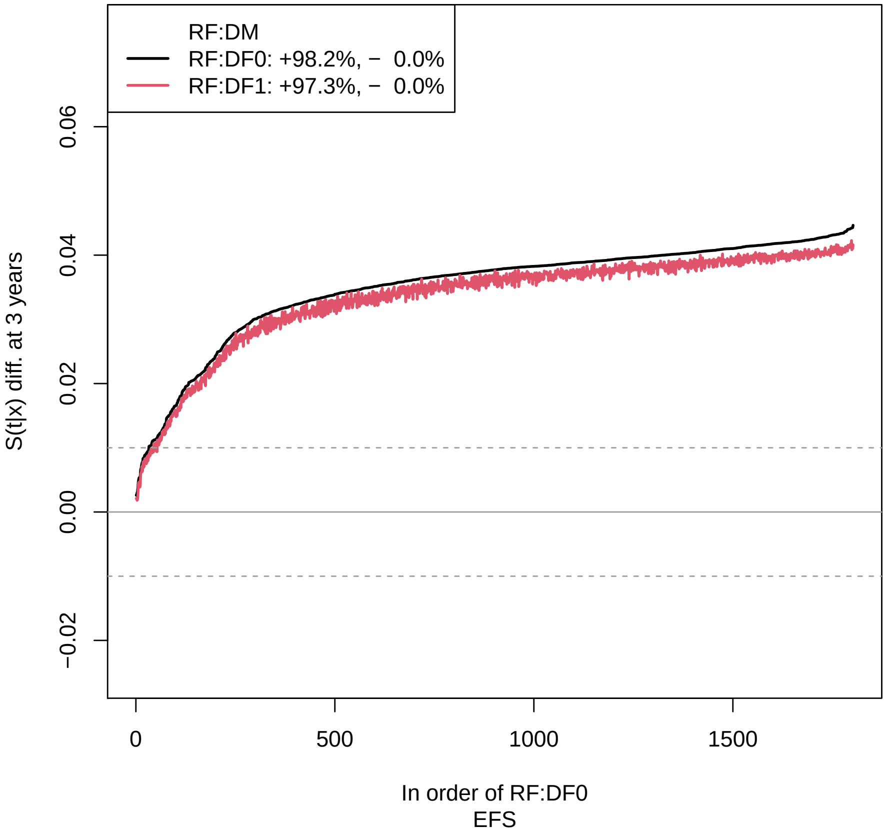


Supplement Figure 8: Waterfall plot of recipient female and donor sex/parity for EFS: validation cohort. Return to cross-reference 9.2.

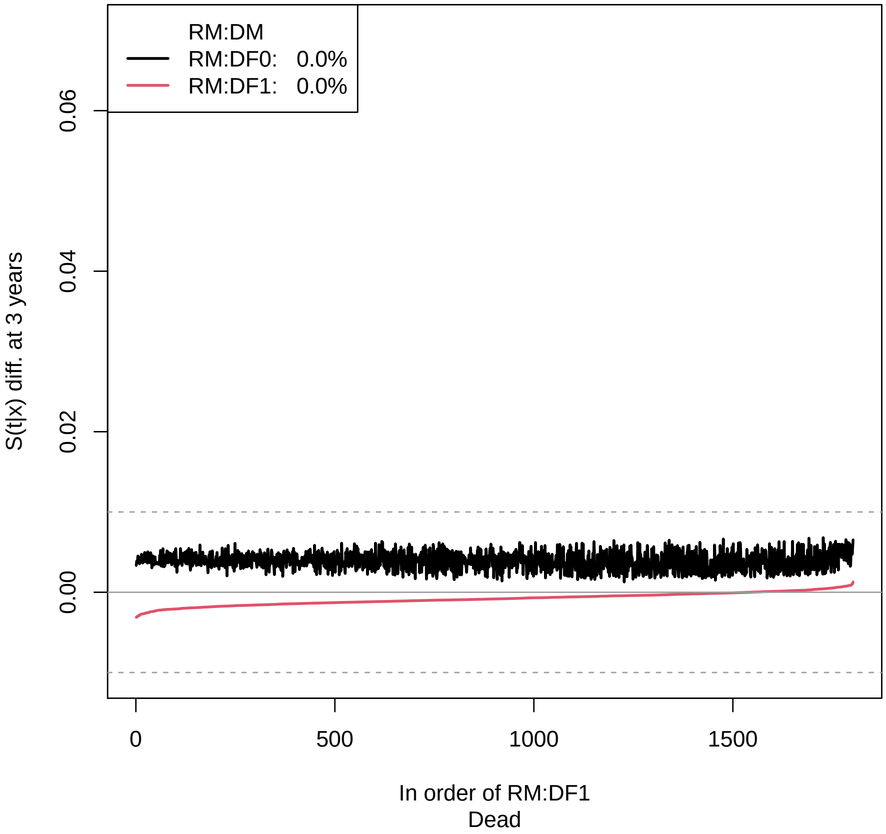


Supplement Figure 9: Waterfall plot of recipient males and donor sex/parity for OS: validation cohort. Return to cross-reference 9.2.

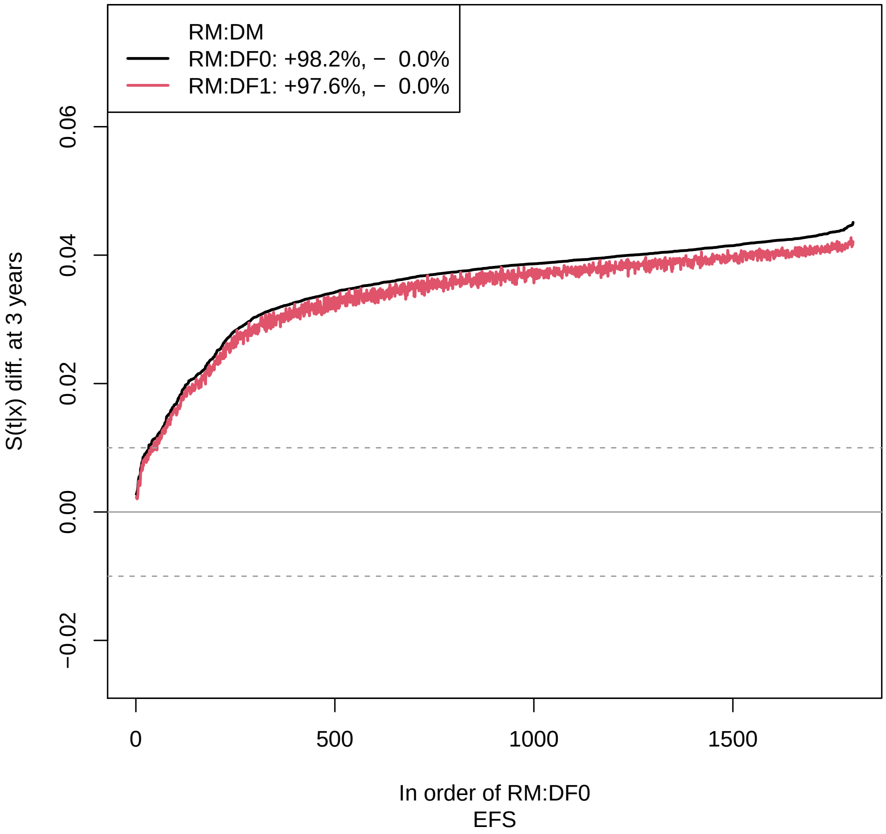


Supplement Figure 10: Waterfall plot of recipient male and donor sex/parity for EFS: validation cohort. Return to cross-reference 9.2.

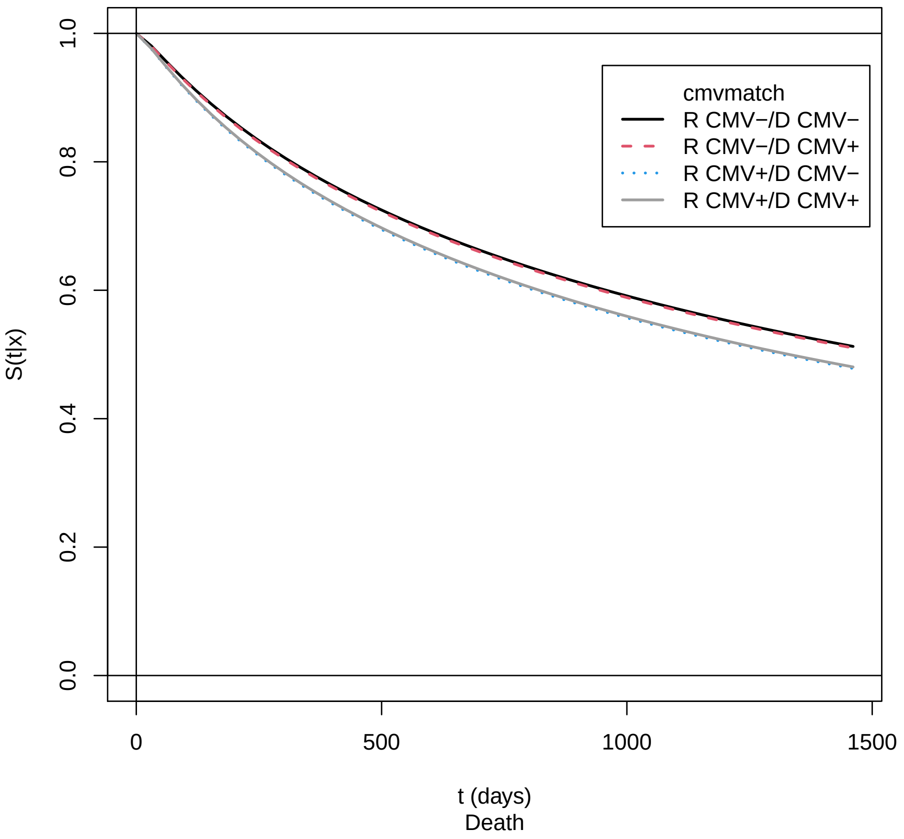


Supplement Figure 11: Marginal effect of CMV match for OS: training cohort. Return to cross-reference 9.2. .

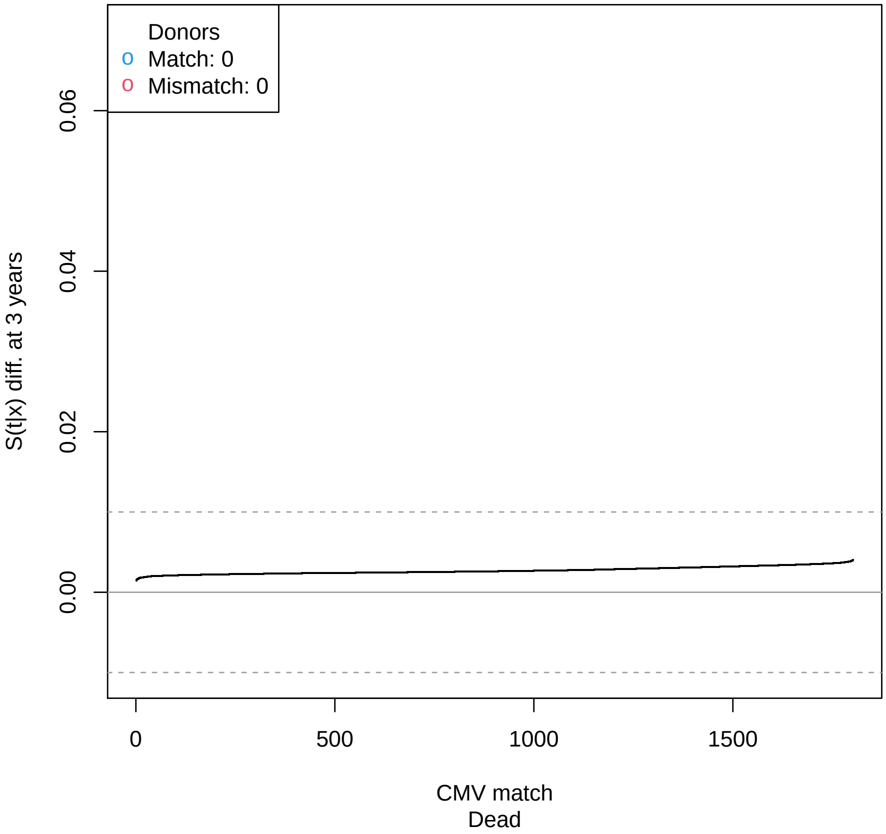


Supplement Figure 12: Waterfall plot of CMV match for OS: validation cohort. Return to cross-reference 9.2.

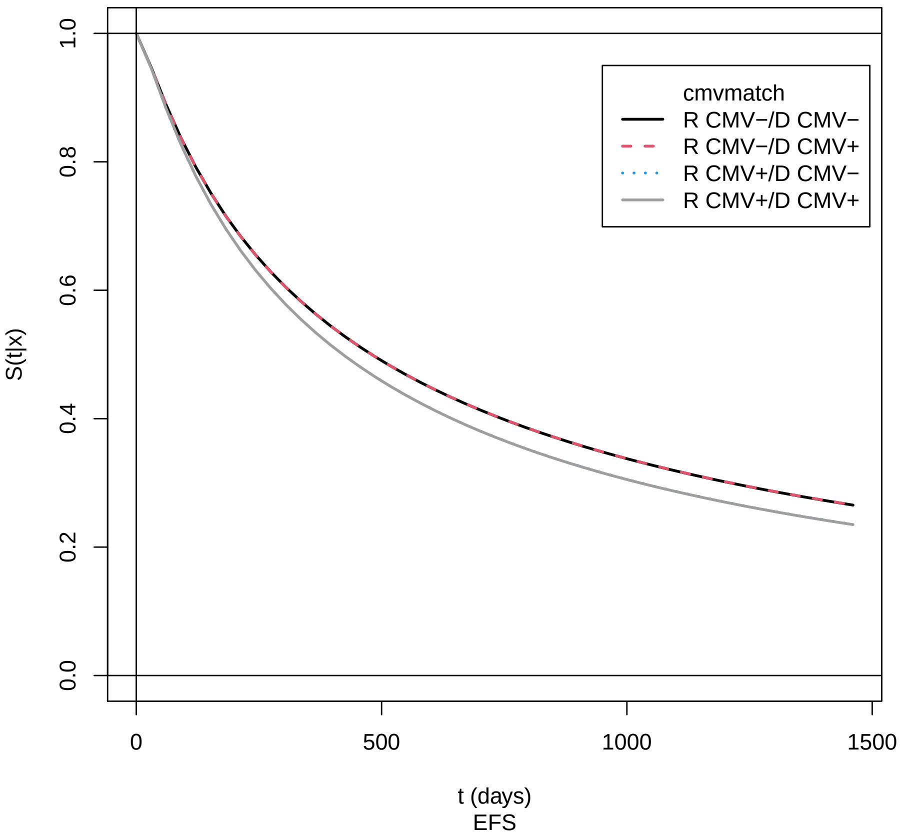


Supplement Figure 13: Marginal effect of CMV match for EFS: training cohort. Return to cross-reference 9.2.

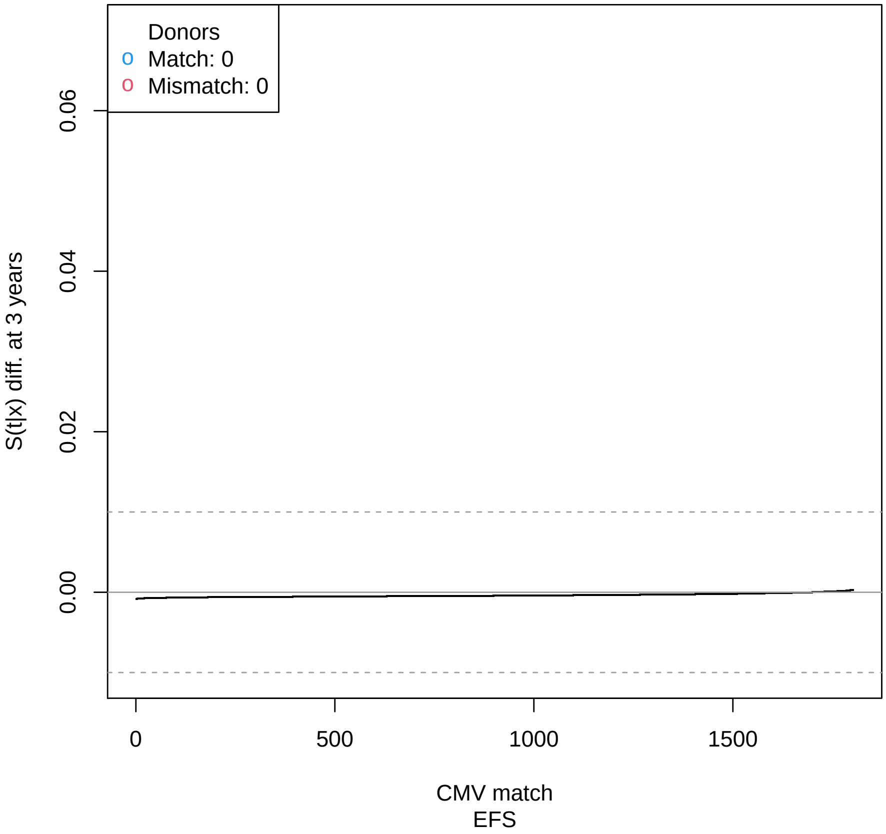


Supplement Figure 14: Waterfall plot of CMV match for EFS: validation cohort. Return to cross-reference 9.2.

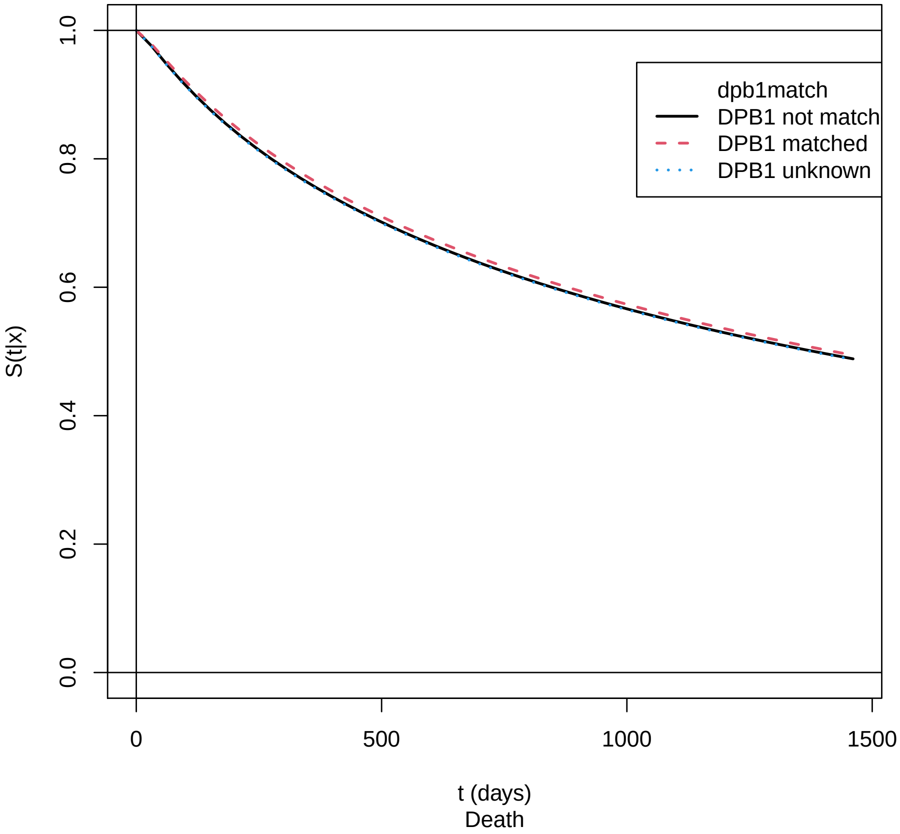


Supplement Figure 15: Marginal effect of DPB1 match/permissive mismatch for OS: training cohort. Return to cross-reference 9.2.

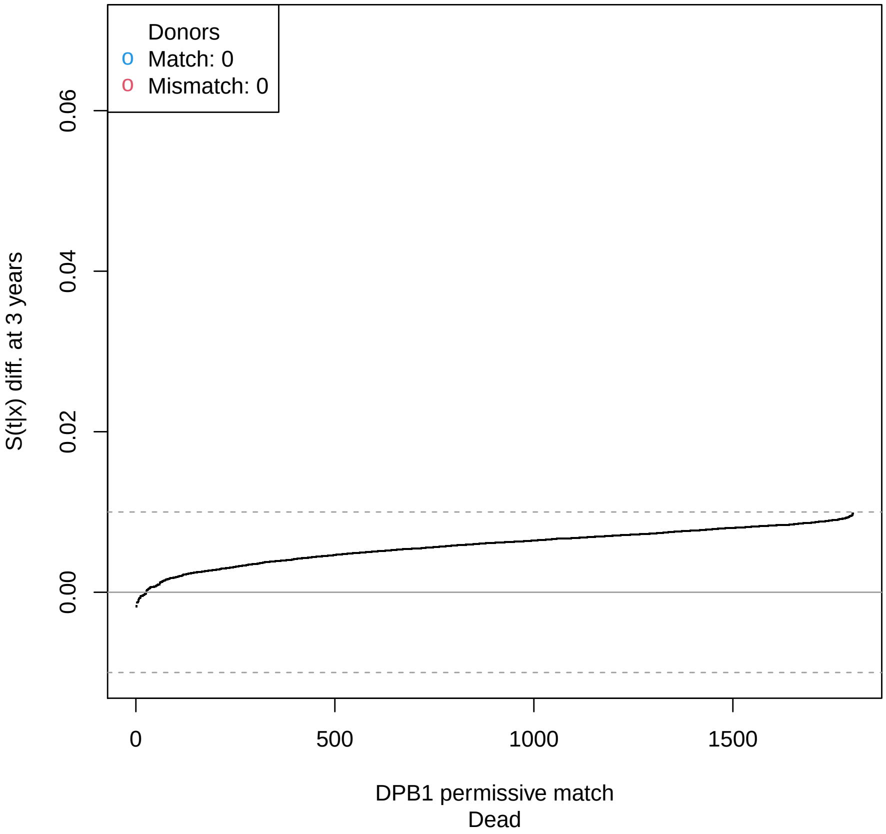


Supplement Figure 16: Waterfall plot of DPB1 match/permissive mismatch for OS: validation cohort. Return to cross-reference 9.2.

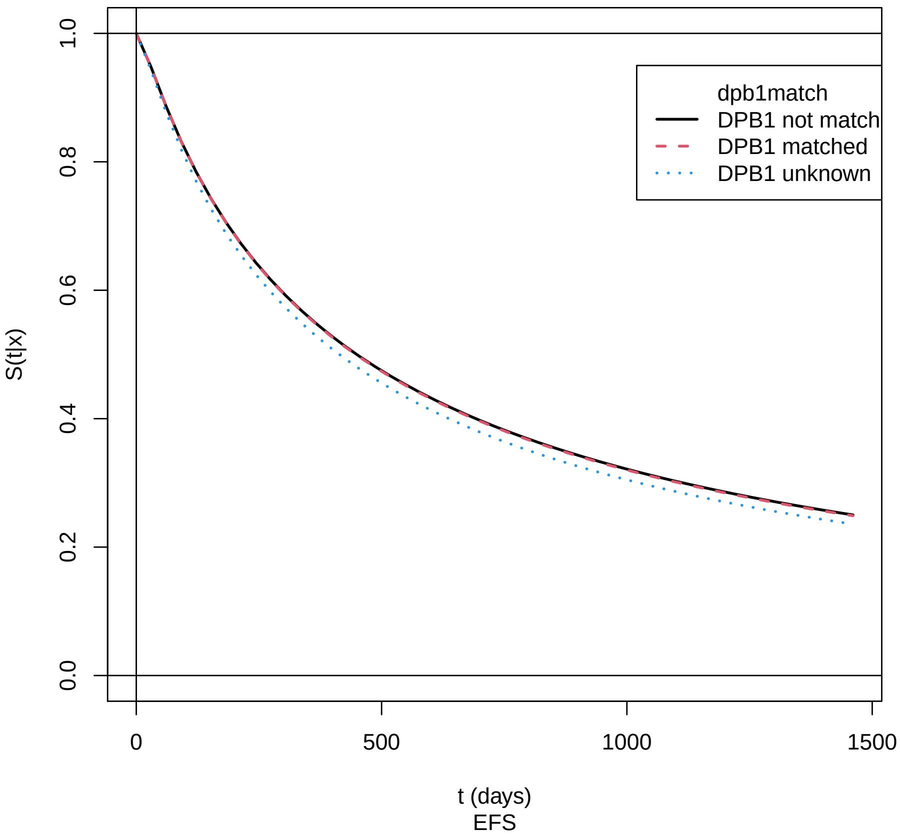


Supplement Figure 17: Marginal effect of DPB1 match/permissive mismatch for EFS: training cohort. Return to cross-reference 9.2.

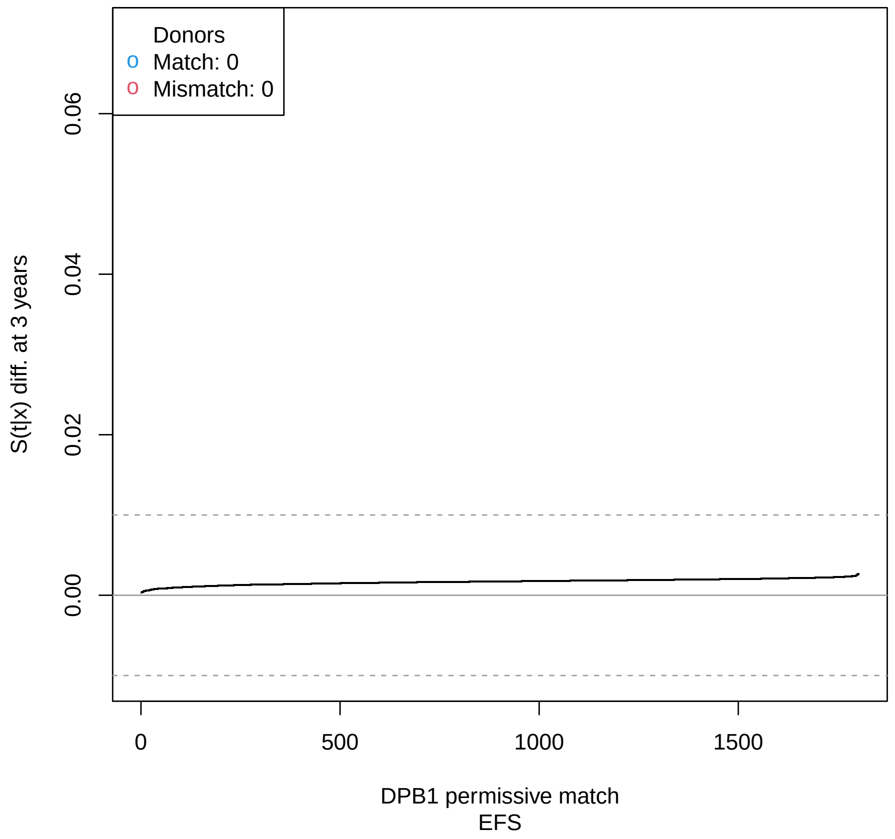


Supplement Figure 18: Waterfall plot of DPB1 match/permissive mismatch for EFS: validation cohort. Return to cross-reference 9.2.

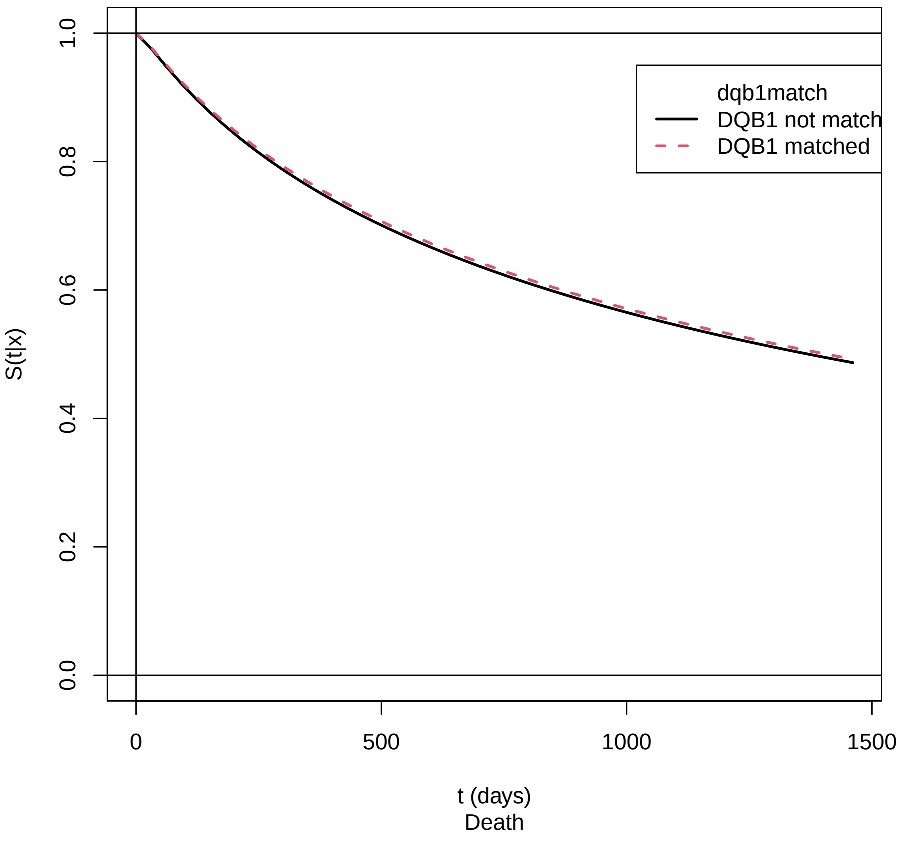


Supplement Figure 19: Marginal effect of DQB1 match for OS: training cohort. Return to cross-reference 9.2.

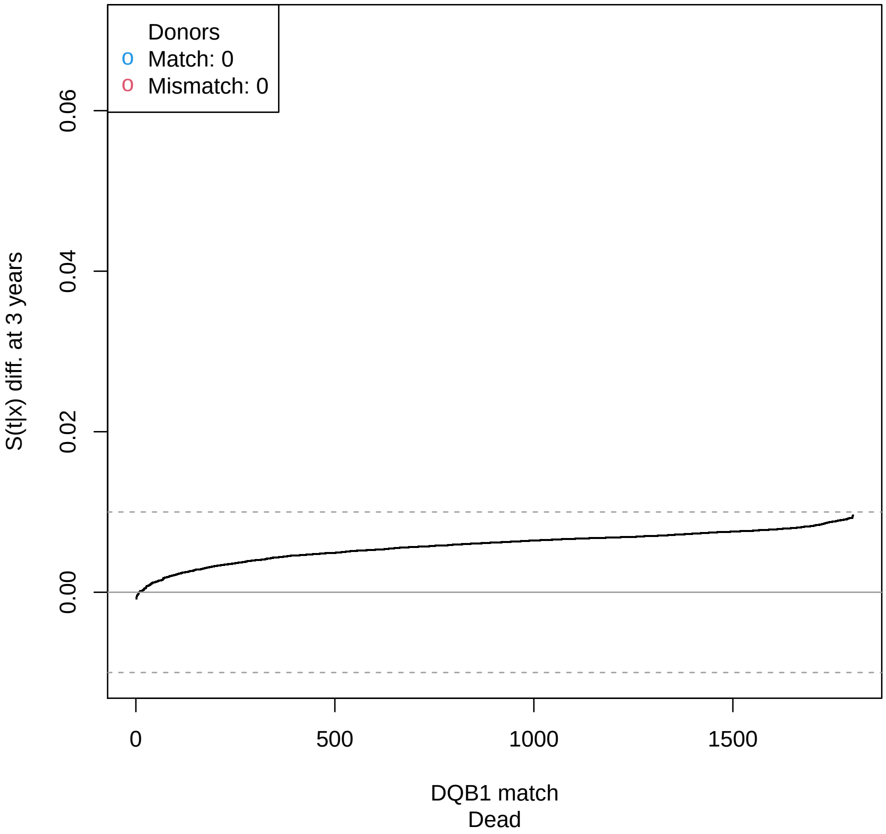


Supplement Figure 20: Waterfall plot of DQB1 match for OS: validation cohort. Return to cross-reference 9.2.

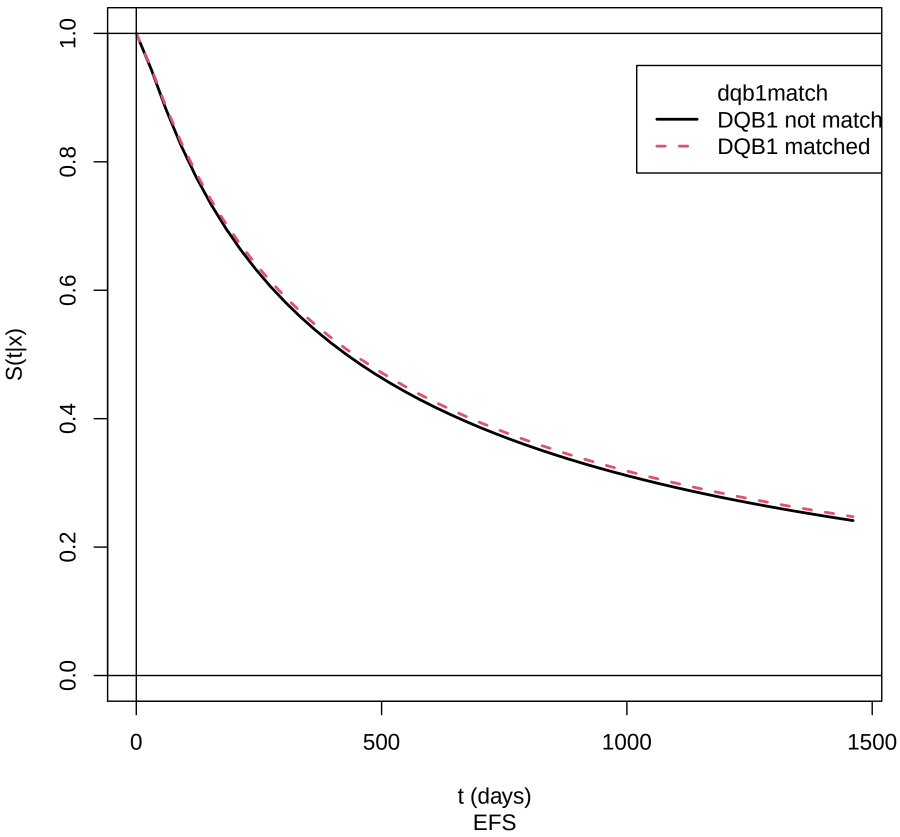


Supplement Figure 21: Marginal effect of DQB1 match for EFS: training cohort. Return to cross-reference 9.2.

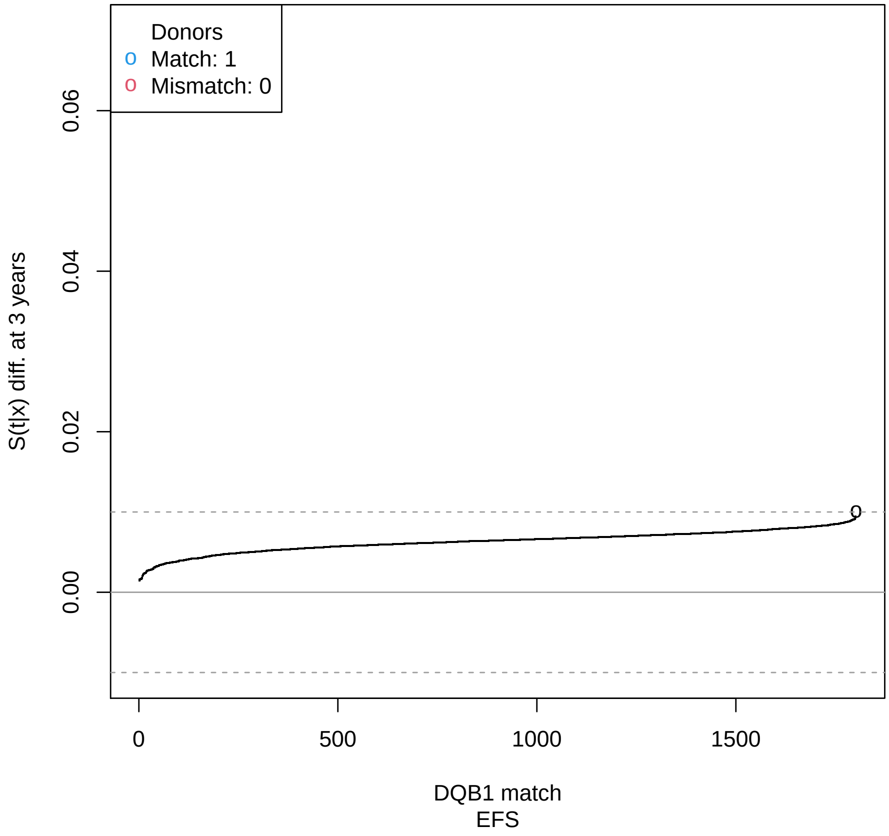


Supplement Figure 22: Waterfall plot of DQB1 match for EFS: validation cohort. Return to cross-reference 9.2.

##### 9.2.2 Figures for recipient characteristics

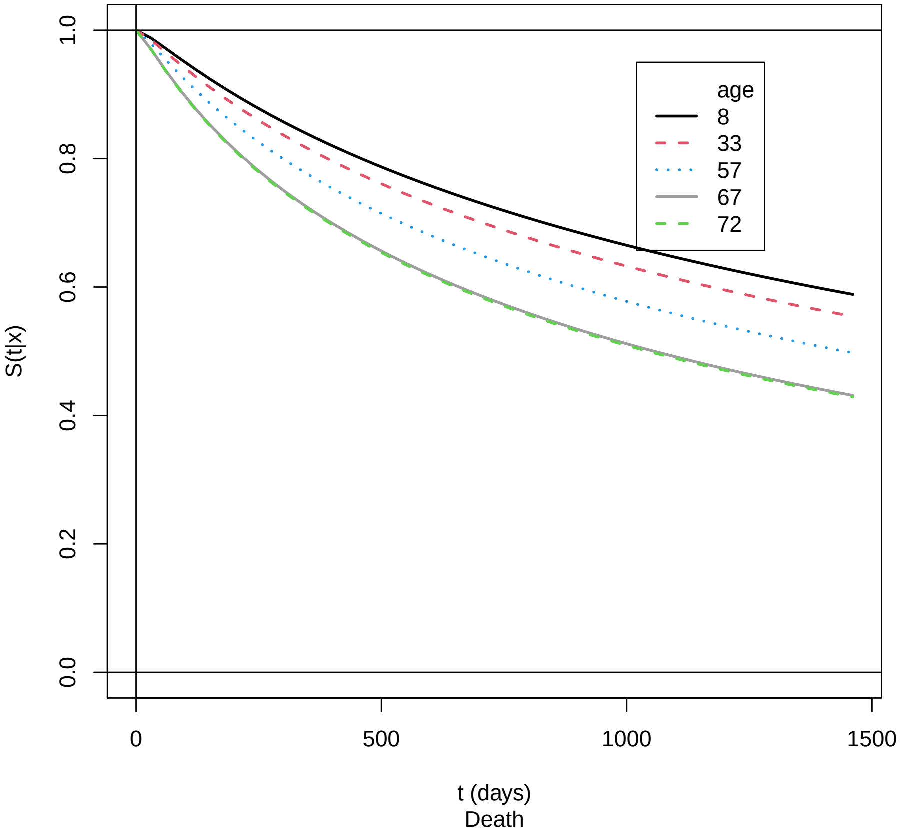


Supplement Figure 23: Marginal effect of recipient age for OS: training cohort. Return to cross-reference 9.2.

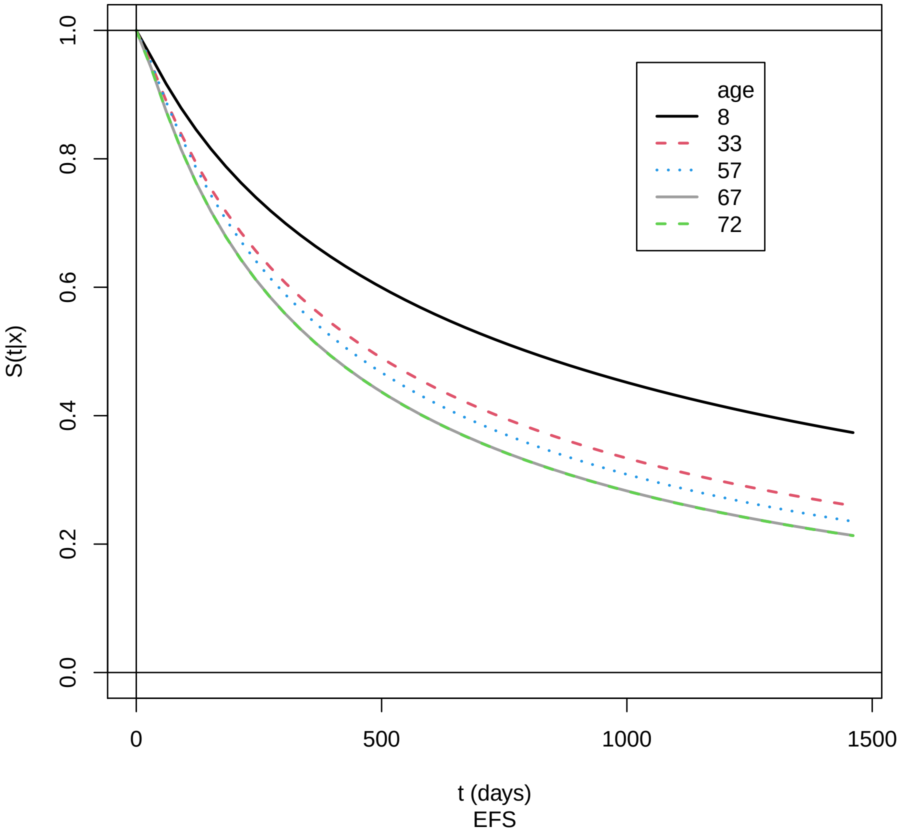


Supplement Figure 24: Marginal effect of recipient age for EFS: training cohort. Return to cross-reference 9.2.

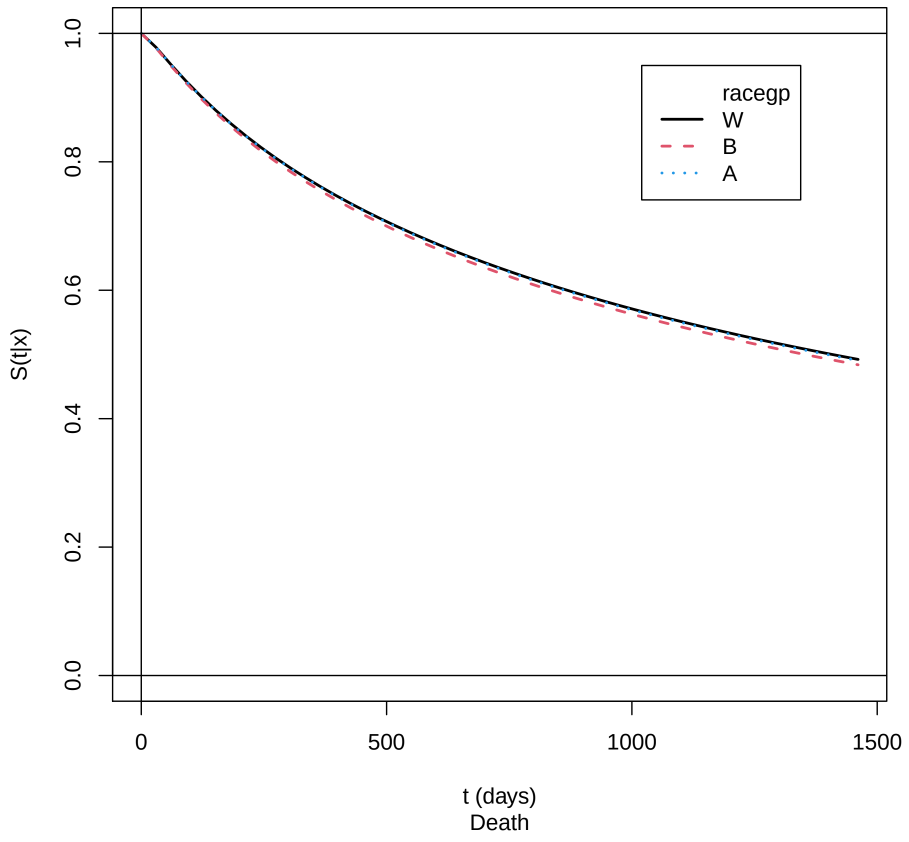


Supplement Figure 25: Marginal effect of race for OS: training cohort. Return to cross-reference 9.2.

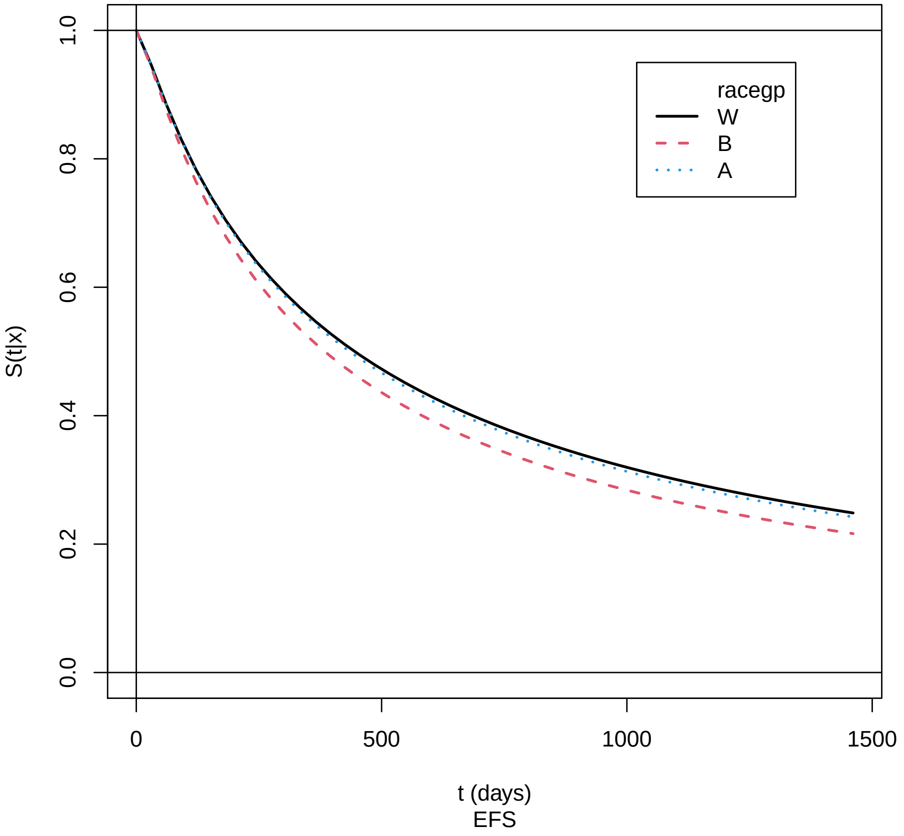


Supplement Figure 26: Marginal effect of race for EFS: training cohort. Return to cross-reference 9.2.

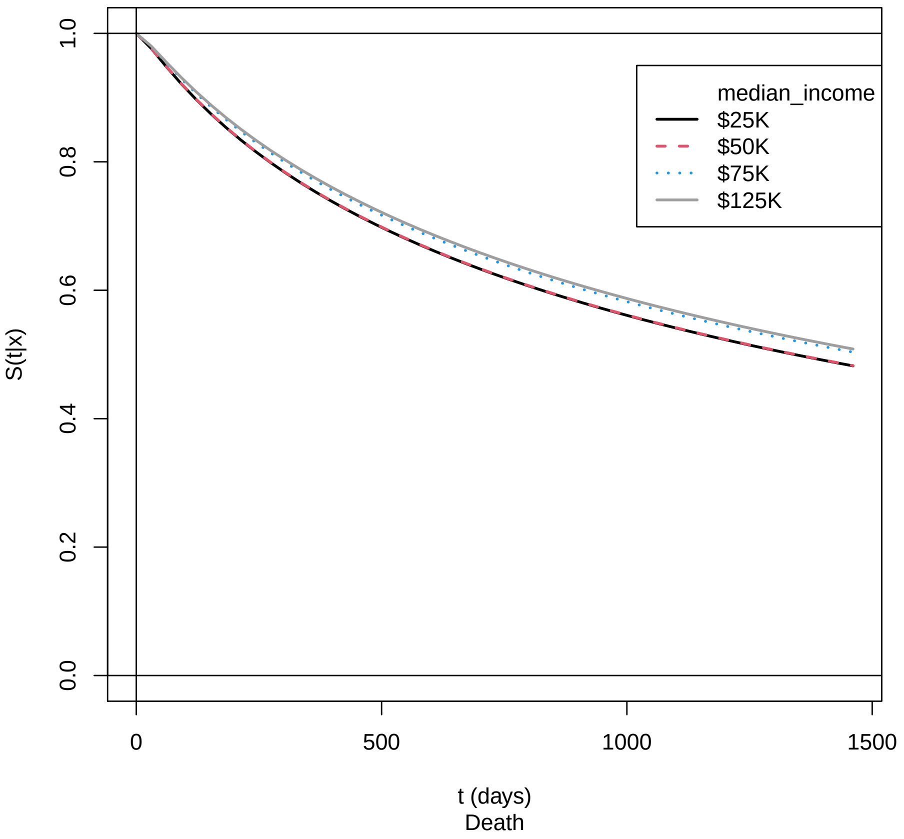


Supplement Figure 27: Marginal effect of ZIP median income for OS: training cohort. Return to cross-reference 9.2.

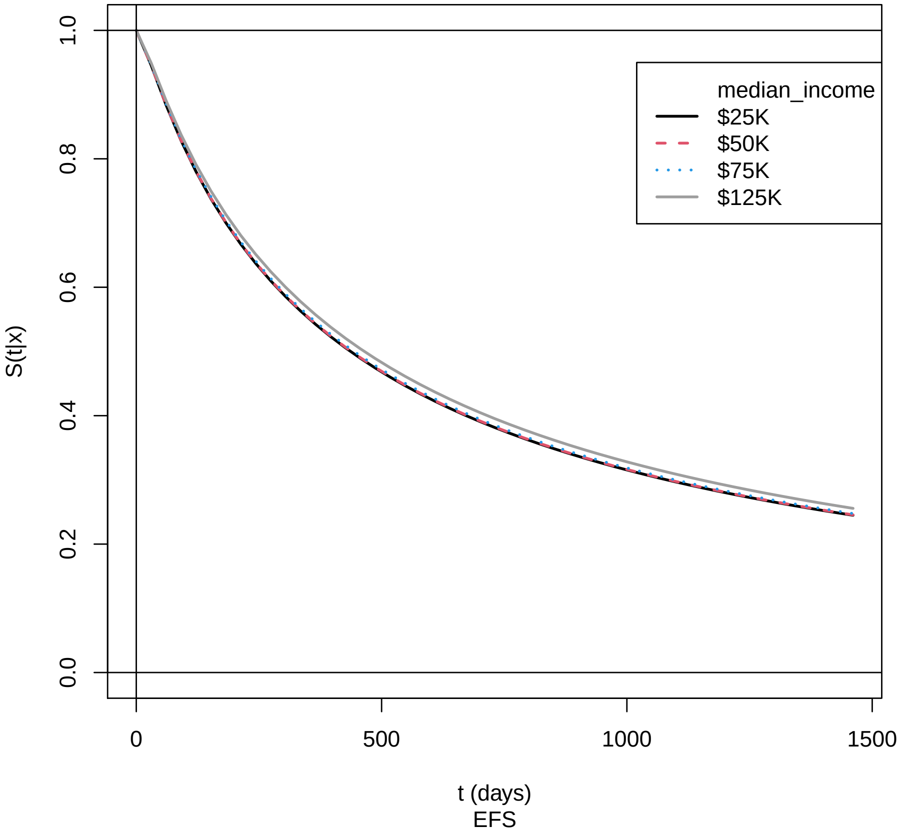


Supplement Figure 28: Marginal effect of ZIP median income for EFS: training cohort. Return to cross-reference 9.2.

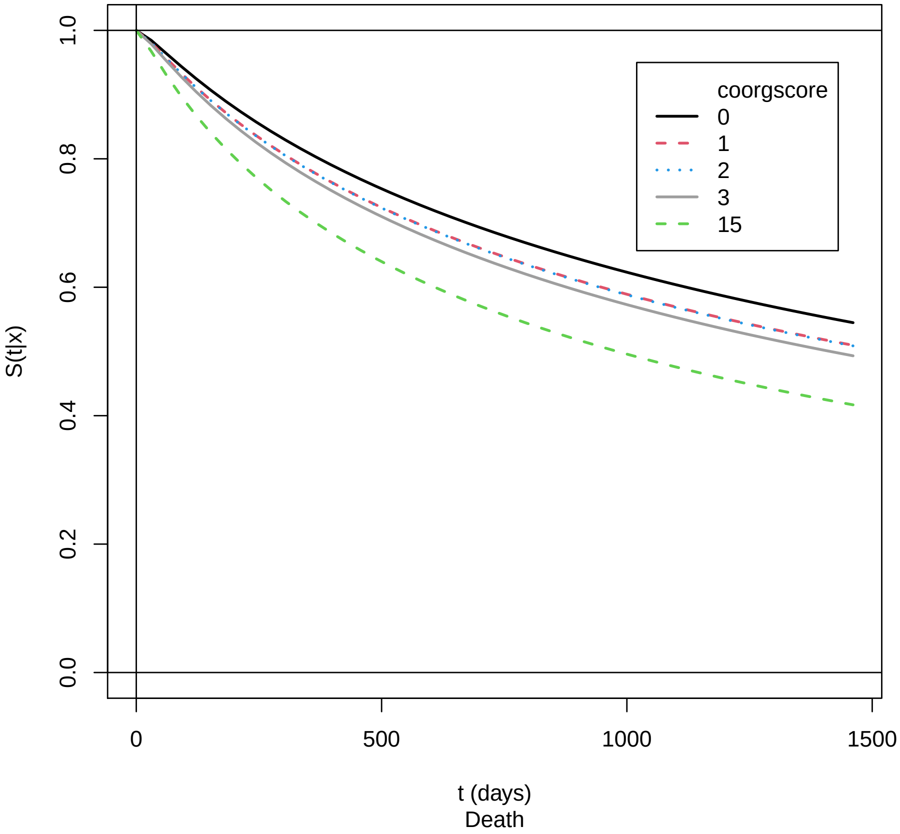


Supplement Figure 29: Marginal effect of HCT-CI for OS: training cohort. Return to cross-reference 9.2.

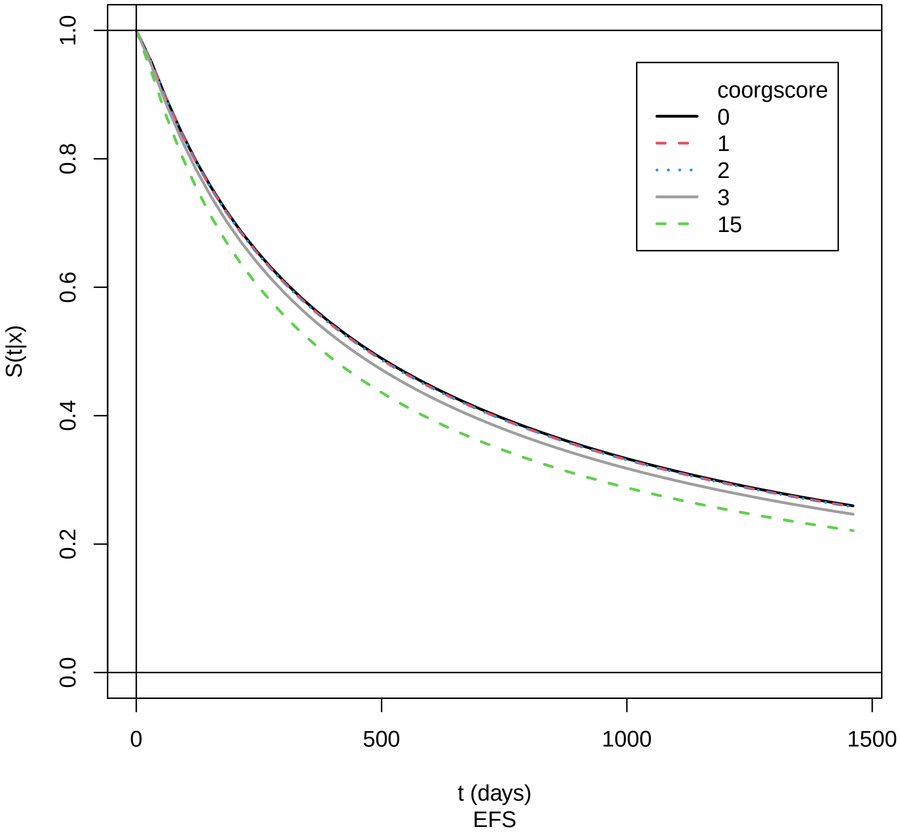


Supplement Figure 30: Marginal effect of HCT-CI for EFS: training cohort. Return to cross-reference 9.2.

##### 9.2.3 Figures for disease/transplant characteristics

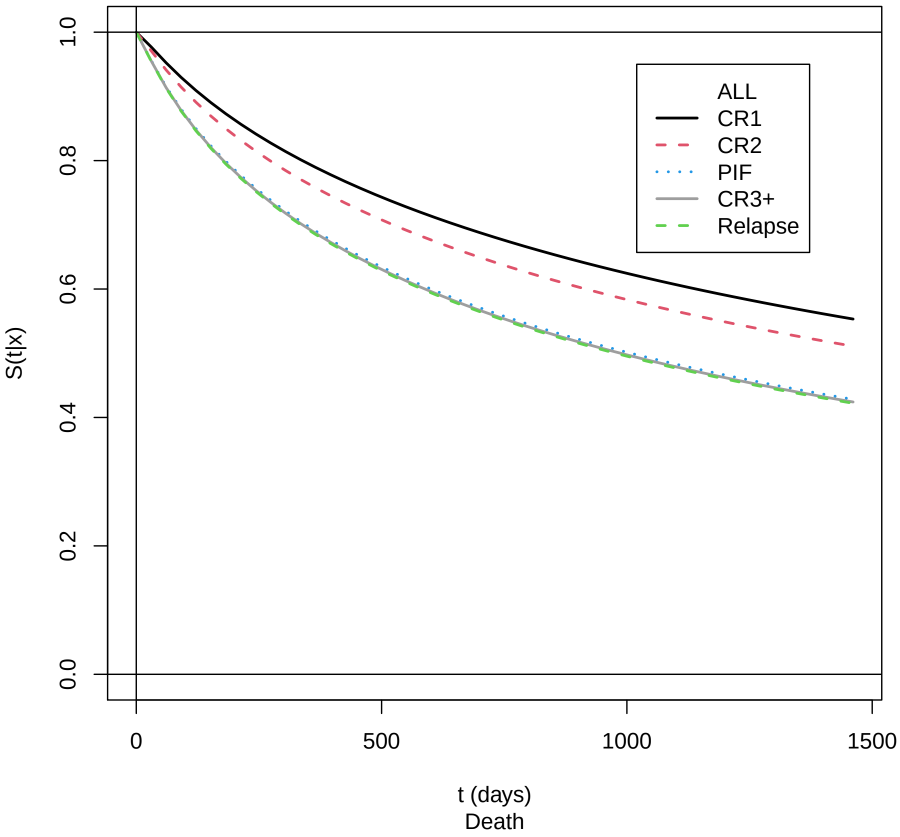


Supplement Figure 31: Marginal effect of ALL status for OS: training cohort. Return to cross-reference 9.2.

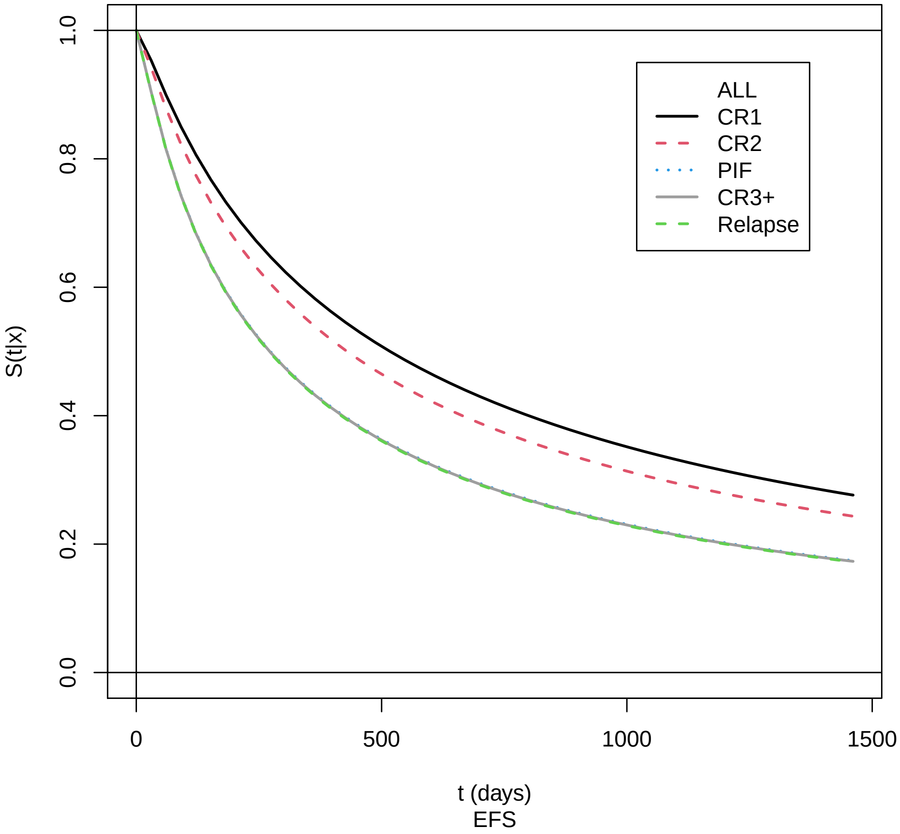


Supplement Figure 32: Marginal effect of ALL status for EFS: training cohort. Return to cross-reference 9.2.

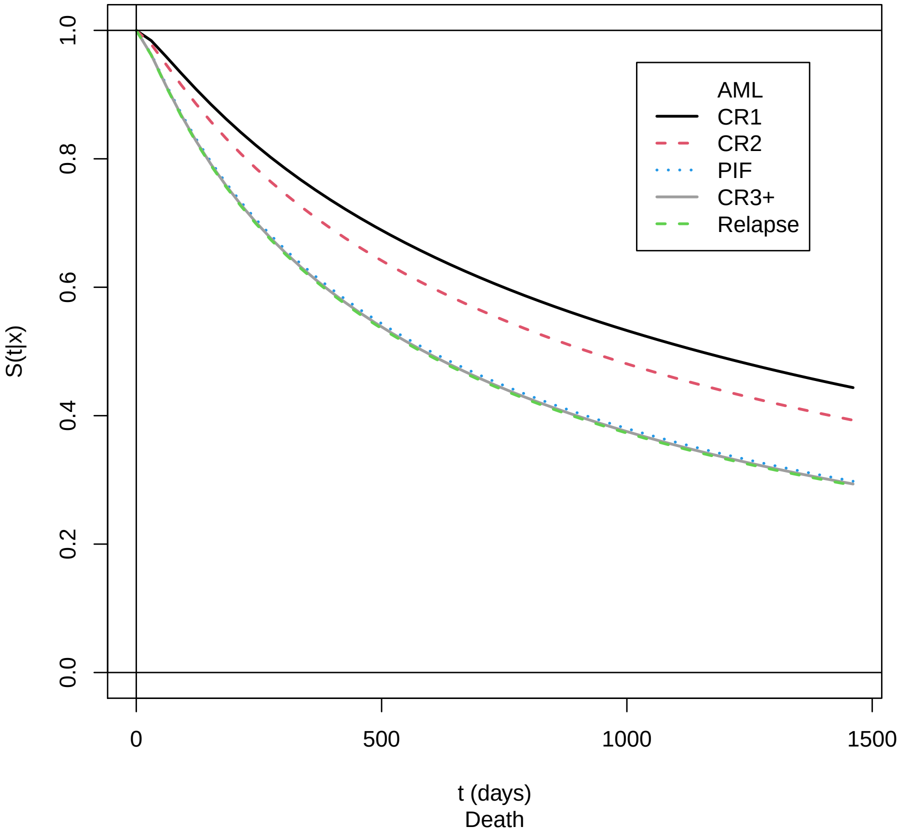


Supplement Figure 33: Marginal effect of AML status for OS: training cohort. Return to cross-reference 9.2.

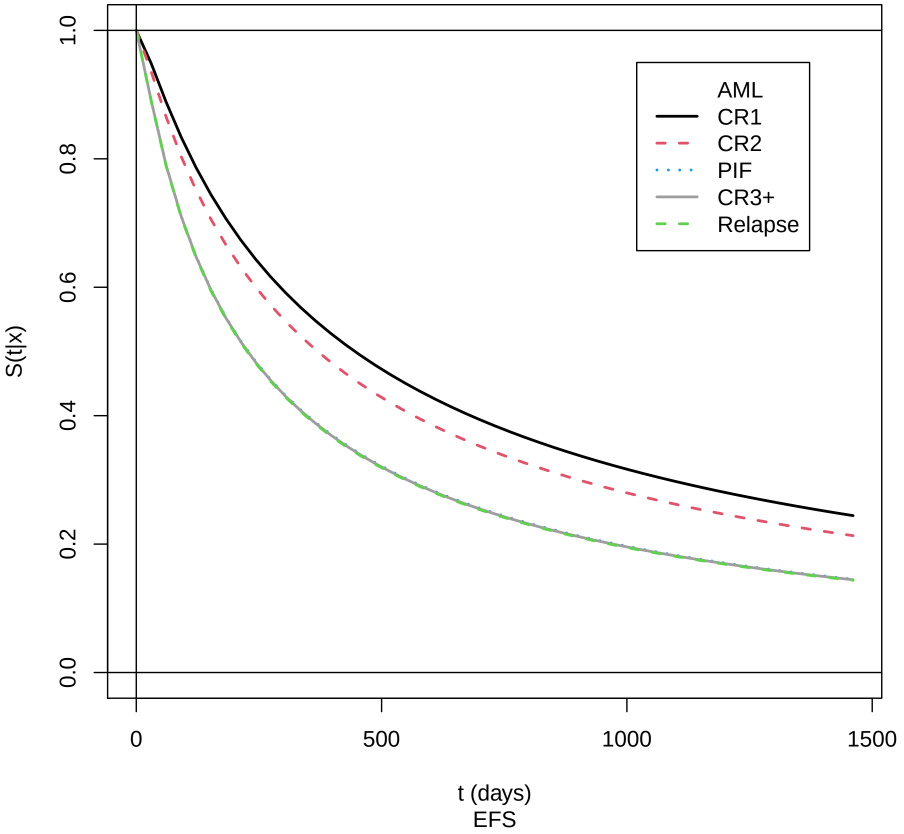


Supplement Figure 34: Marginal effect of AML status for EFS: training cohort. Return to cross-reference 9.2.

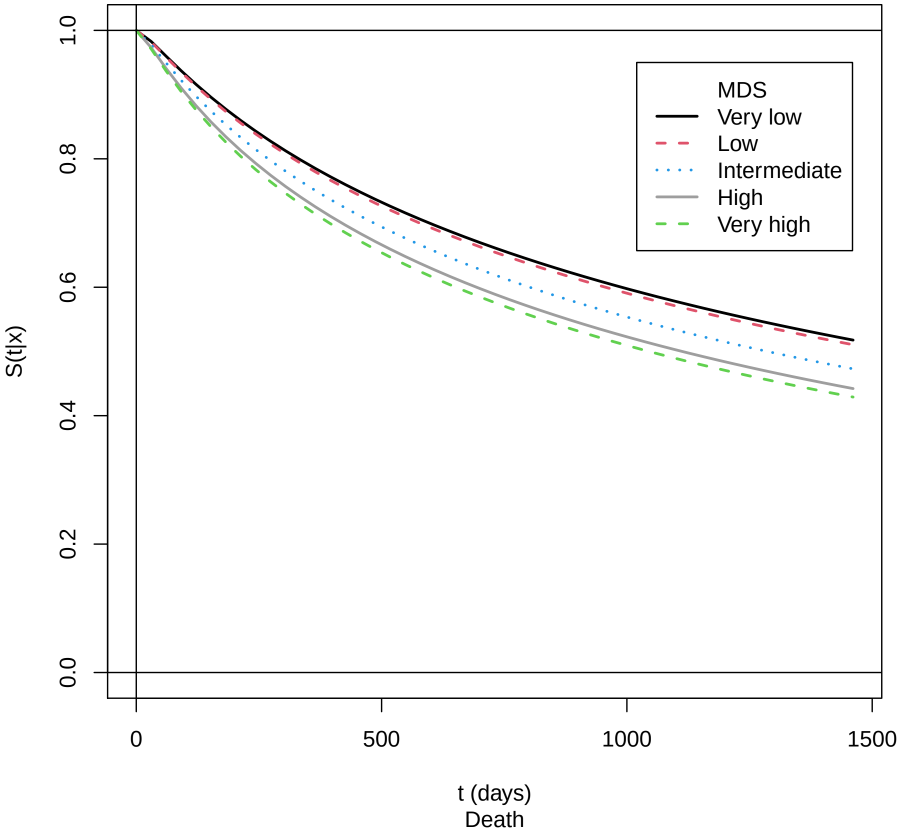


Supplement Figure 35: Marginal effect of MDS status for OS: training cohort. Return to cross-reference 9.2.

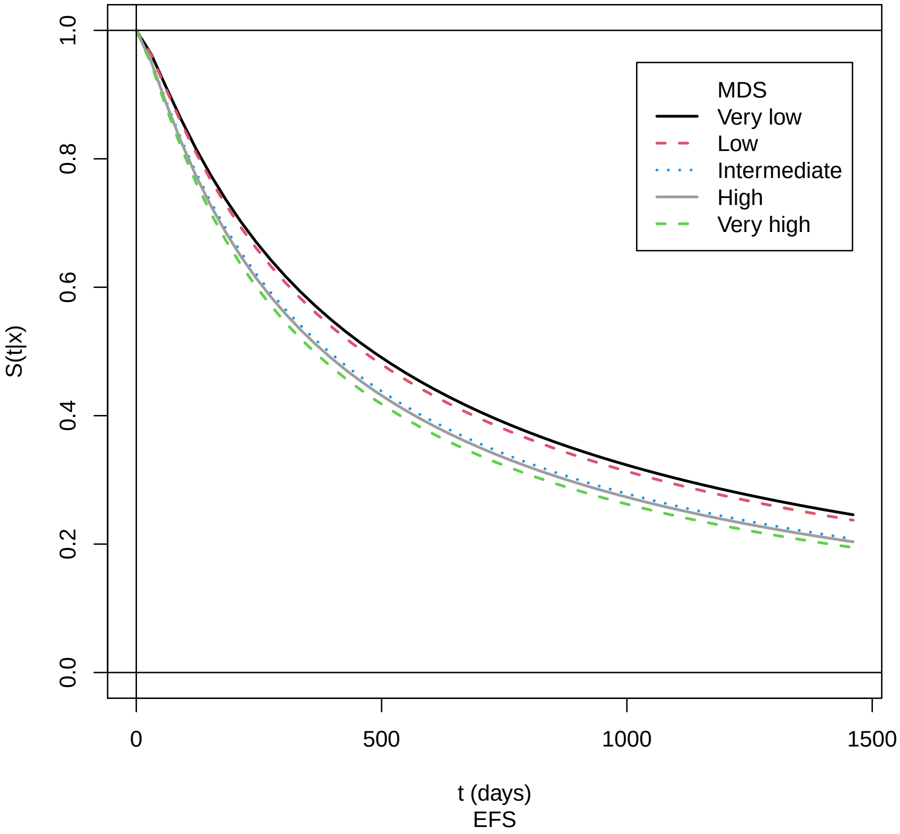


Supplement Figure 36: Marginal effect of MDS status for EFS: training cohort. Return to cross-reference 9.2.

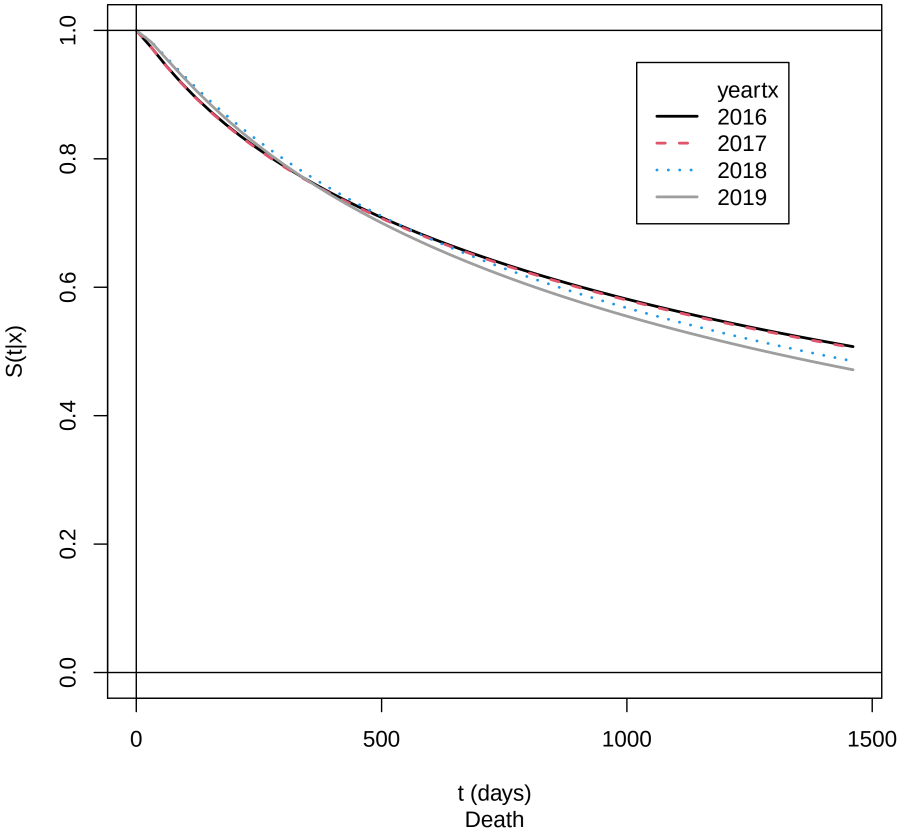


Supplement Figure 37: Marginal effect of transplant year for OS: training cohort. Return to cross-reference 9.2.

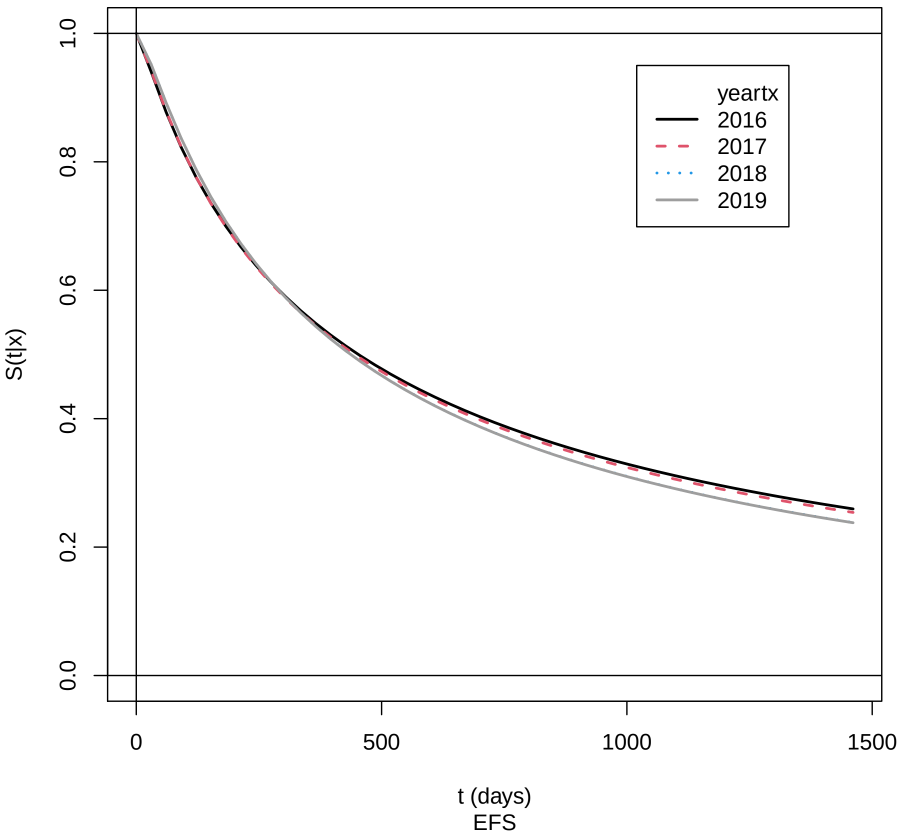


Supplement Figure 38: Marginal effect of transplant year for EFS: training cohort. Return to cross-reference 9.2.
